# Multi-omics data integration from patients with carotid stenosis illuminates key molecular signatures of atherosclerotic instability

**DOI:** 10.1101/2025.05.12.25327328

**Authors:** Vivek Das, Sampath Narayanan, Xiang Zhang, Otto Bergman, Djordje Djordjevic, Malin Kronqvist, Melody Chemaly, Glykeria Karadimou, Sofija Vuckovic, Inika Prasad, Andrew J. Buckler, Karin Conde Knape, Natasha Barascuk Michaelsen, Ulf Hedin, Ljubica Matic

## Abstract

**Background:** Understanding the pathophysiology of unstable atherosclerosis is imperative to prevent myocardial infarction and stroke. We used multi-omics integration to identify key molecular targets with diagnostic and therapeutic potential.

**Methods:** Biobank of Karolinska Endarterectomies encompassing patients with symptomatic (S) and asymptomatic (AS) carotid atherosclerosis, was the main resource. Plaques, peripheral blood monocytes and plasma sampled locally from around plaque or periphery of n>700 individuals, were profiled by transcriptomics, proteomics and metabolomics. A supervised feature-selection method DIABLO was used for *per* patient data integration. Multi-omics layers were integrated separately across local and peripheral disease sites, and their intersection, with stratification for symptomatology. Identified analytes were investigated using scRNAseq, clinical and outcome data.

**Results:** In peripheral circulation, FABP4, IL6, Bilirubin and Sphingomyelin were the most prominent analytes. F11, ANGPTL3, ICOSLG, ITGB1 and Sphingomyelin were enriched in the local disease site, while FABP4, C1R, IL6, Bilirubin and Sphingomyelin appeared at the intersection. Coagulation, necroptosis, inflammation and cholesterol metabolism were confirmed as key pathways determining symptomatology. Clinical analyses showed an impact of lipid-lowering therapy on ICOSLG expression, anti-hypertensives on plasma FABP4 and BLVRB levels, anti-diabetics on plasma Sphingomyelins, while no medications affected ANGPTL3. Association with future adverse events was shown for plasma Bilirubin, Sphingomyelin, ANGPTL3 and ICOSLG plaque levels. Open-source target genetic analyses suggested causal involvement of F11, C1S, EGFR, IL6, ANGPTL3 in the disease.

**Conclusions:** Using an innovative, deep-data framework, this study provides confirmatory and novel information on mechanisms behind atherosclerotic instability. The findings raise possibilities for translational prioritizations to aid personalized medicine.

**Structured Graphical Abstract:** 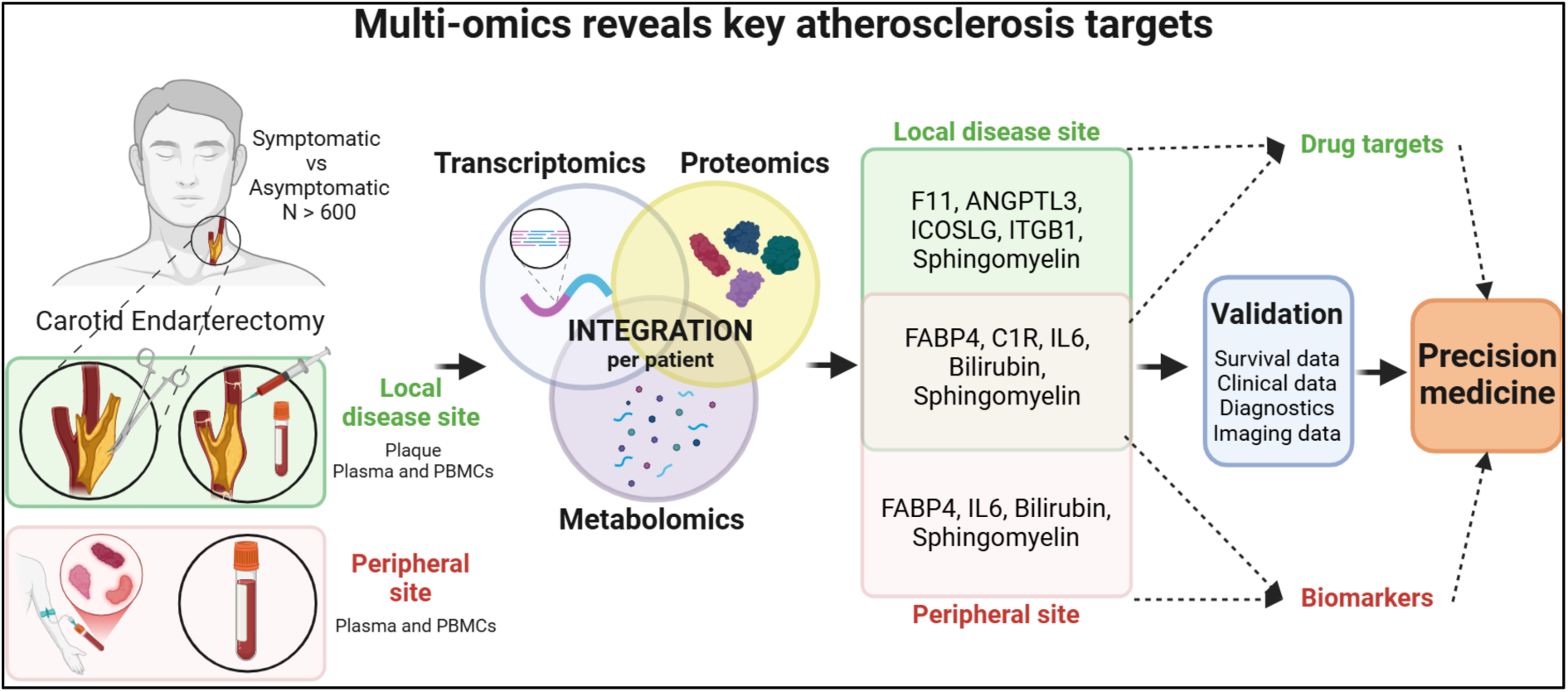

**Key Question:** This study performed first-of-a-kind, orthogonal, per-patient multi-omics integration from a large carotid stenosis biobank, with an aim to identify key molecular signatures and pathways of human atherosclerotic instability.

**Key Finding:** The complex multi-omics design coupled with deep-data analyses, enabled the discovery of numerous confirmatory and novel molecular signatures implicated in patient symptomatology. Extended validation analyses elucidated their cellular sources, associations with plaque morphology, clinical biochemistry, medication and long-term patient outcomes.

**Take-home Message:** The findings are interesting for further investigation with respect to druggable targeting or plasma biomarkers, altogether leading to improved patient phenotyping and precision medicine potential in cardiovascular disease.

## Introduction

Despite the profound impact of atherosclerotic cardiovascular disease (ASCVD) on global mortality and morbidity from myocardial infarction (MI) or ischemic stroke (IS), drug development in this area has plateaued in recent years. Based on the Framingham Heart Study^1^, ASCVD research has for decades focused on pathophysiological pathways related to various risk factors, resulting in the development of successful preventive strategies targeting lipids, thrombosis, diabetes, hypertension and lately also inflammation. Nevertheless, there is a considerable residual risk with 5-10% optimally-managed patients experiencing a major cardio- or cerebro-vascular event (MACCE) three years after the initial event and a life-time risk of up to 50-60%^2^. More specifically, asymptomatic (AS, stable) carotid disease is associated with an annual stroke risk of 1-3%, whereas symptomatic (S, unstable) patients with either transient ischemic attack (TIA) or minor stroke (MS) have a more than 10-fold increased risk^1^. While current prevention strategies have clearly curbed mortality rates, substantial risk remains and importantly, existing treatments lack specificity for diverse patient subgroups. Thus, there is a need to subcategorize patients, where precise, individualized therapeutic strategies grounded in causal vascular disease biology can be applied.

Atherogenesis is critically associated with hyperlipidaemia, infiltration and retention of ApoB containing lipoproteins in the artery wall, triggering inflammatory responses with endothelial dysfunction and subendothelial infiltration of monocytes, which develop into foam cells and form fatty streaks^3, 4,5^. Cytokines and chemokines secreted by macrophages over time promote recruitment of other inflammatory cells^5^, which gradually leads also to the activation of medial smooth muscle cells (SMCs) into a secretory and replicating phenotype that engage in intimal remodelling and fibrous cap formation^6, 7^. Progressive inflammation contributes to plaque instability by inhibiting collagen production, stimulating neovessel formation and intraplaque hemorrhage, as well as matrix metalloproteinases that degrade the fibrous cap, which altogether may lead to its rupture, atherothrombosis and fatal clinical manifestations^8 9^.

In previous years, analyses of human atherosclerosis from single-omics, i.e. transcriptomic or proteomic datasets, combined with both patient and plaque phenotyping, have facilitated discoveries of novel molecular targets and pathways in plaque instability^10, 11^. More recently, advanced large-scale studies have mapped genetic risk loci across diverse populations^12–14^, performed in-depth molecular subtyping^13, 15^ and dissected cell-specific disease mechanisms^6, 16–23^, resulting in identification of several subclinical phenotypes of ASCVD^24^. Especially single-cell RNA sequencing (scRNAseq) has improved the characterization of different cell populations in atherosclerosis, with identification of various T-cells, macrophages and SMCs subsets. Moreover, single-cell atlases of ASCVD have revealed that, in addition to atherogenic dyslipidemia and systemic inflammation, a considerable portion of genetic risk can be explained by the dysfunction of endothelial cells and SMCs^17, 25–27^, raising a notion that cells of the vessel wall could be a source of novel biomarkers or therapeutic targets.

However, few holistic systems biology approaches have been applied in the field so far, although efforts to elucidate complex disease-underlying mechanisms by integrating different layers of omics information using machine learning technologies have shown promise in other areas^28^. To this end, DIABLO (Data Integration Analysis for Biomarker discovery using Latent cOmponents) has been utilized, as a novel integrative method that seeks for common information across different high-dimensional data types. DIABLO is based on pairwise linear combinations of factors that reduces dimensionality of the data, captures biological variability and identifies correlated molecular analytes associated with categorical phenotypes. It has been shown that DIABLO identifies features with superior biological relevance, achieving predictive performance that supersedes other state-of-the-art approaches^29^.

In this study, our objective was to accelerate the identification of novel biomarker and therapeutic candidates specifically targeting instability in late-stage atherosclerosis. To this end, we aimed to leverage the power of multi-omics integrations from a large human biobank using DIABLO, never previously attempted on this scale and complexity. Novel bioinformatic workflows for multi-dimensional disease assessment and target identification were developed by combining global transcriptomic, proteomic and metabolomic profiling of matched plaques, PBMCs and plasma from patients undergoing stroke-preventive carotid endarterectomy (CEA), utilizing the Biobank of Karolinska Endarterectomies (BiKE)^30^. The synergy of orthogonal omics datasets analyzed here was shown to be necessary for unravelling the deep mechanistic insights that underly heterogeneity in S *vs.* AS patients, as well as locally around the lesion *vs.* in peripheral circulation. The generated results illuminate both classical and novel aspects of ASCVD and enable a more comprehensive understanding of this patient population.

## Materials and Methods

Detailed Material and Methods are provided in the **Supplementary File**.

## Results

### Study design and workflow

Biomaterial from atherosclerosis patients consisted of matched plaques, PBMCs and peripheral as well as “local” plasma samples, retrieved in the proximity of the lesion. The study was conducted in 3 major stages **(Figure 1)**, all stratified by comparisons between S *vs.* AS patients in order to identify molecular signatures of atherosclerotic instability. The first stage consisted of large-scale single-omics data analyses (plaque and PBMCs transcriptomics, proteomics and metabolomics of plasma sampled form peripheral circulation or locally from the disease site). In the second stage, multi-omics was designed to integrate 3 combinations of datasets separately, where Combination 1 integrated data from peripheral circulation to identify potential biomarkers of atherosclerotic instability; Combination 2 integrated data from the local disease site, previously shown to be enriched with lesion-derived analytes^11^; and Combination 3 integrated data from plaques and peripheral circulation with an aim to detect molecular signatures in blood that originate from the culprit lesion. In the third stage, targeted translational analyses were performed for the top candidates from multi-omics integrations, associating their plaque and/or plasma levels with cell types, clinical biochemistry parameters, medication and outcomes in patients. **Supplementary Figure I** shows the data generated for this study.

**Figure 1:**
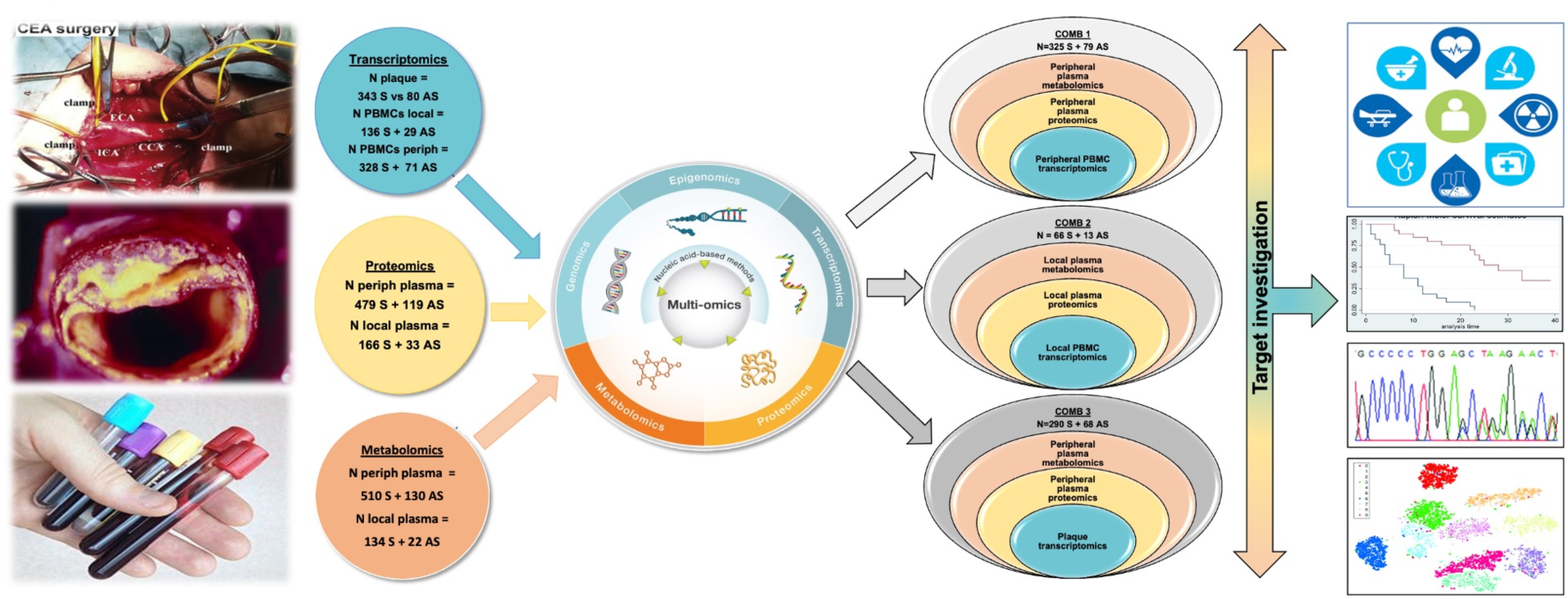
Illustration of the study design and multi-omics integration workflow. Samples for the Biobank of Karolinska Endarterectomy (BiKE) were successively collected at carotid endarterectomy (CEA) surgery since 2002, covering the whole region of Stockholm, Sweden. Plaques, PBMCs and plasma were profiled using large-scale, state-of-the-art omics technologies with transcriptomics by RNAseq, proteomics by OLINK and metabolomics by LC-MS/MS. Single-omics analyses were performed in each layer, comparing samples from symptomatic *vs*. asymptomatic patients. Multi-omics integration by DIABLO tool was performed on matched data from patients, coupling orthogonally peripheral (Combination 1) or local (Combination 2) sites, as well as an intersection of both (Combination 3). Top analytes from each Combination passing the correction for multiple comparisons, were investigated further on individual level by interrogation of clinical, epidemiological, follow-up, genetic data from the same patients as well as public scRNAseq datasets and database mining.

### Single-omics analyses comparing S vs. AS patients

#### Transcriptomics reveals enrichment of epithelial-to-mesenchymal transition, angiogenesis, inflammation and cholesterol metabolism in plaques and PBMCs from S vs. AS patients

After quality control, analysis was performed for n>16 000 transcripts in a total of n=423 plaques (343 S + 80 AS patients), n=165 local PBMCs (136 S + 29 AS patients) and n=399 peripheral PBMCs (328 S + 71 AS patients) **(Supplementary Figure II)**. Differential transcript level analyses were performed comparing S *vs.* AS patients, with sex and age as covariates and correction for multiple comparisons, in PBMCs and plaque samples separately. This analysis resulted in 152 differentially regulated transcripts in plaques, 9 in local and 586 in peripheral PBMCs.

The most significantly upregulated genes in plaques from S patients were i.e. CHI3L2, CXCL6, CLEC4G, MMP16, CFB and PI15 **(Supplementary Table I)**. Gene set enrichment analysis showed that epithelial-to-mesenchymal transition, TNFa and IL6 signalling, IFNg response, hypoxia and angiogenesis, ECM reorganisation and bone remodelling, cholesterol homeostasis and glycolysis were some of the most important pathways in plaques. Functional associations among the significantly dysregulated genes were related to cell migration, tissue remodelling, actin binding. Most active associations were found at the basement membrane and actomyosin cytoskeleton, while interactions enriched in the network were related to focal adhesions and ECM **(Supplementary Figure III)**.

Similar analyses in local PBMCs showed an upregulation of i.e. CCL2, GPNMB, MMP19, TREM2, IL10 and an enrichment of epithelial-to-mesenchymal transition, TNFa and IL6 signalling, IFNg response, coagulation, heme metabolism and angiogenesis, cholesterol homeostasis and glycolysis as important pathways **(Supplementary Table II, Supplementary Figure III)**.

In peripheral PBMCs 102 transcripts were upregulated, such as LGMN, IGFBP4, SYNPO, PODXL, COL6A1, while again epithelial-to-mesenchymal transition, TGFB/WNT and IL2 signalling, coagulation, angiogenesis, cholesterol homeostasis and cell apoptosis processes were enriched. Functional interactions among the differentially regulated genes in both local and peripheral PBMCs included cartilage development and proteoglycans **(Supplementary Table III, Supplementary Figure III)**.

#### Proteomics highlights enrichment of chemokine signalling in both local and peripheral plasma, while IL17 pathway was specifically upregulated locally

After quality control, analysis was performed for n=442 proteins in a total of n=598 peripheral (479 S + 119 AS patients) and n=199 local plasma samples (166 S + 33 AS patients) **(Supplementary Figure II)**. Differential protein levels analysis was performed comparing S *vs.* AS patients, with sex and age as covariates, in local and peripheral samples separately. After adjustments, 68 proteins were found to be dysregulated in peripheral and 36 in local plasma **(Supplementary Tables IV and V)**, of which n=15 proteins were shared between the local and peripheral plasma **(Supplementary Figure IV).** Commonly dysregulated proteins included known atherosclerosis markers (i.e. CSF1, CDH5, FASLG, NOTCH3, IL6, IL6RN, EGFR, CXCL8, PLAUR), but also some less explored candidates (i.e. ICOSLG, PCOLCE, FUT5, CCL20, CA6, KITLG).

Enrichment analysis confirmed the overall upregulation of chemokine signalling and cytokine-receptor interactions in both local and peripheral plasma, while IL17 pathway was specifically enriched locally **(Supplementary Figure IV)**. Functional network coupling reinforced the notion that several of these proteins were also key drivers of the differential changes observed in both local and peripheral blood (IL6, EGFR, CXCL8), while others had a more prominent function in local (CSF1) or peripheral (TIMP1, CXCL12) disease sites, respectively **(Supplementary Figure IV)**.

#### Metabolomics shows upregulation of Sphingomyelins and downregulation of Bilirubin degradation products in association with symptomatology

After quality control, analysis was performed for n=1011 metabolites in a total of n=640 peripheral (510 S + 130 AS patients) and n=156 local plasma samples (134 S + 22 AS patients) **(Supplementary Figure II)**. Differential metabolite analysis was performed comparing S *vs.* AS patients, with sex and age as covariates, in local and peripheral plasma samples separately. In peripheral plasma, there were n=213 metabolites that were found to be nominally dysregulated and in the local plasma n=76 **(Supplementary Tables VI and VII).** Totally n=31 metabolites were commonly dysregulated in both local and peripheral plasma **(Supplementary Figure V).** Notably, tryptophan betaine, Bilirubin and Bilirubin degradation products were downregulated, while various Sphingomyelins, glyco-beta-muricholate and mannonate were upregulated in S *vs.* AS local and peripheral plasma comparisons.

Enrichment analysis confirmed these results, reinforcing the increase in Sphingomyelins in both local and peripheral blood, while Bilirubin degradation products remained significantly repressed in peripheral blood **(Supplementary Figure V)**.

### Multi-omics integration analyses

Multi-omics integration of BiKE datasets was next performed with a rationale to identify complex orthogonal relationships across molecular species/analytes associated with unstable carotid atherosclerosis. To this end, matched patient samples and analytes for each of the 3 multi-omics combinations (peripheral circulation, local disease site, intersection between plaque and peripheral site) were first identified **(Supplementary Figures VI and VII).**

#### Multi-omics data integration in peripheral circulation reveals FABP4, Sphingomyelins, IL6 and Bilirubin degradation as key signatures associated with symptoms

Contribution of various metabolites, proteins and transcripts from relevant single-omics layers (peripheral PBMC transcriptomics + peripheral plasma proteomics + peripheral plasma metabolomics, referred to as Combination 1) to the integrated multi-omics model in peripheral circulation was analyzed using the DIABLO tool and stratified according to symptomatology **(Figure 2A)**. The analysis showed that a signature composed of *i.e.* upregulation of various Sphingomyelins and downregulation of Bilirubin degradation metabolites, along with upregulation of FABP4, IL6, ANG, TNFRSF11 and SPP1 proteins, as well as upregulation of ARG2, SPAG8 with downregulation of MMEL1, OSBP, LGSN transcripts, was collectively enriched in the peripheral circulation of S patients. Joint enrichment analysis of all 3 omics layers in the multi-omics model, showed an enrichment of hematopoietic cell lineage pathways, arginine metabolism and inflammation, while Ras and PI3K/Akt signalling were repressed **(Figure 2B)**. Correlation based network analysis highlighted an association of FABP4 with various Sphingomyelins and Bilirubin degradation, while evidence-based network centered around IL6 interactome and Bilirubin degradation products **(Figure 2C)**.

**Figure 2:**
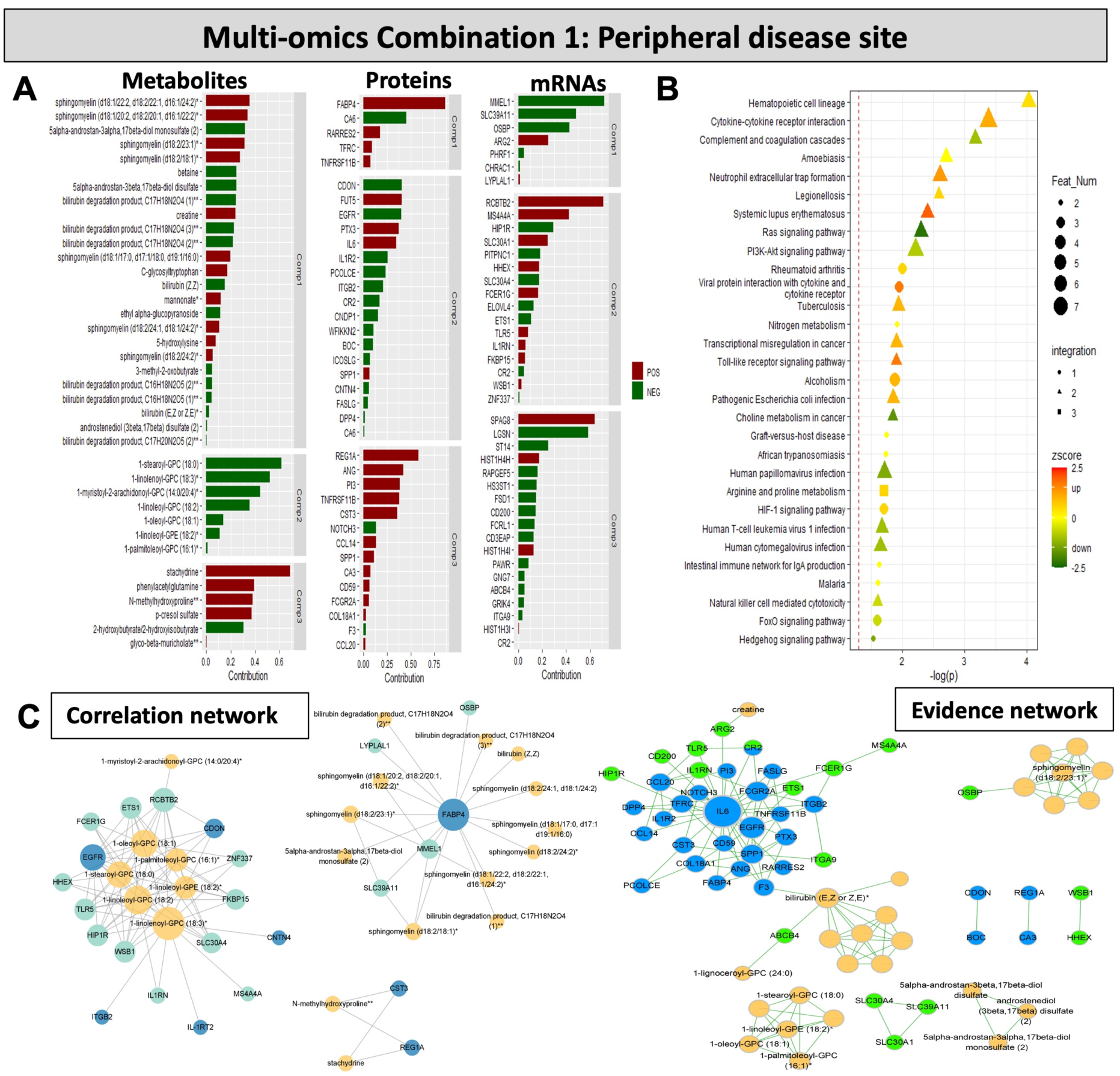
Multi-omics data integration around the peripheral disease site (Combination 1). **A)** Contribution of various metabolites, proteins and mRNAs from relevant single-omics layers to the integrated multi-omics model with 3 components. Bars represent the contribution of each feature to the symptomatic patient phenotype. Direction of contribution indicated with color (red-positive, green-negative). **B)** Joint enrichment analysis of all 3 omics layers in the multi-omics model. Circle indicates that the enriched term contains only one type of omics data, triangle represents terms containing 2 omics types, and square for 3 types. Direction of effect indicated with colour (green-to-red, legend). **C)** Correlation based network (rho >0.5) (to the left) and evidence-based network (to the right) of different analytical features from the multi-omics analyses. Color represents the type of omics data (blue, green and yellow referring to proteins, transcripts and metabolites respectively), size of the node represents the degree of significance.

#### Multi-omics data integration at the local disease site reveals Sphingomyelins, ANGPTL3, F11, ICOSLG, ITGB1, SOX11, MPZL1, TFPI as key signatures associated with symptoms

Next, the contribution of various analytes from 3 omics layers to the integrated multi-omics model around the local disease site (local PBMC transcriptomics + local plasma proteomics + local plasma metabolomics, Combination 2) was also analyzed in connection to the presence of symptoms **(Figure 3A)**. Again, the analysis highlighted the strong upregulation of various Sphingomyelins and showed that a signature composed from *i.e.* Sphingomyelin metabolites, along with the dysregulation of F11, ST6GAL1, ANGPTL3, ICOSLG, ITGB1 proteins, as well as dysregulation of SOX11, MPZL1, TFPI, PTGR1 transcripts, was collectively enriched at the local disease site in S patients. Joint enrichment analysis of omics layers in this model, demonstrated coagulation, cholesterol metabolism and necroptosis as the most prominent pathways **(Figure 3B)**. Correlation based network analysis highlighted a direct association of F11 and ANGPTL3 with various Sphingomyelins and cholesterol, while evidence-based network centered around the ITGB1, F11 and ANGPTL3 interactomes **(Figure 3C)**.

**Figure 3:**
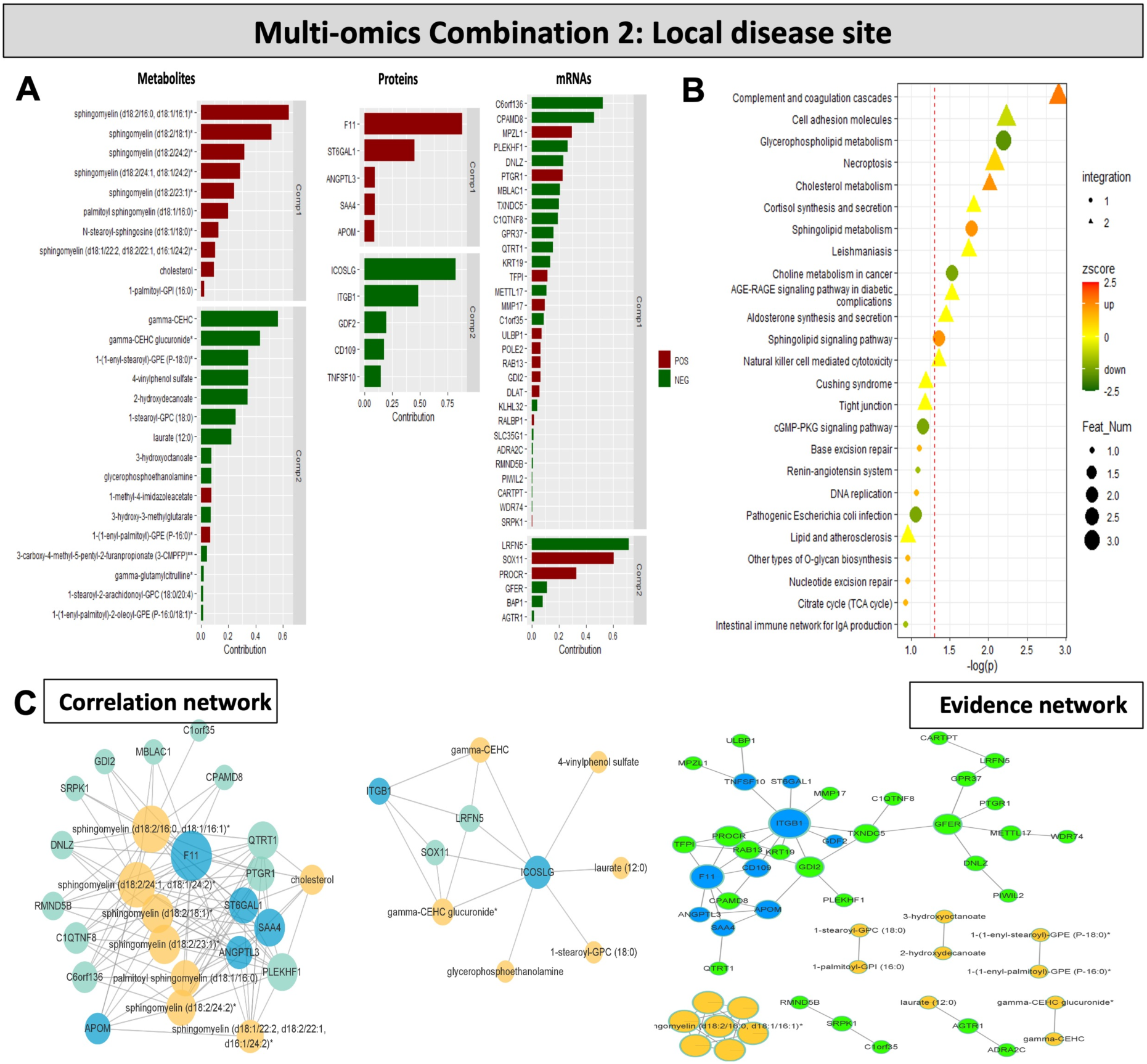
Multi-omics data integration around the local disease site (Combination 2). **A)** Contribution of various metabolites, proteins and mRNAs from relevant single-omics layers to the integrated multi-omics model with 2 components. Bars represent the contribution of each feature to the symptomatic patient phenotype. Direction of contribution indicated with color (red-positive, green-negative). **B)** Joint enrichment analysis of all 3 omics layers in the multi-omics model. Circle indicates that the enriched term contains only one type of omics data, triangle represents terms containing 2 omics types, and square for 3 types. Direction of effect indicated with colour (green-to-red, legend). **C)** Correlation based network (rho >0.5) (to the left) and evidence-based network (to the right) of different analytical features from the multi-omics analyses. Color represents the type of omics data (blue, green and yellow referring to proteins, transcripts and metabolites respectively), size of the node represents the degree of significance.

#### Multi-omics data integration combining plaque and peripheral disease site reveals Sphingomyelins, fatty acids, Bilirubin degradation products, complement components, FABP4 and IL6 as key signatures associated with symptoms

Finally, the contribution of various analytes from relevant single-omics layers to the integrated multi-omics model at the intersection between peripheral and local disease site (plaque transcriptomics + peripheral plasma proteomics + peripheral plasma metabolomics, Combination 3) was analyzed and stratified by symptomatology **(Figure 4A)**. Once again, the analysis showed that a signature composed from *i.e.* upregulation of various Sphingomyelins and downregulation of Bilirubin degradation metabolites and many fatty acids, along with the upregulation of RARRES2, FABP4, PLAUR, IL6 and downregulation of PCOLCE, COMP, ICOSLG proteins, as well as upregulation of TNC, MARCO, C1R, C1RL, C1S, CSB transcripts, was enriched in S patients. Joint enrichment analysis of all 3 omics layers in the multi-omics model, highlighted various infectious pathways **(Figure 4B)**. A correlation-based analysis highlighted two main networks: one *via* a direct FABP4 association with RARRES2 and various Bilirubin degradation products, and another *via* several complement components (C1S, C1R, CFB) and various fatty acids. An evidence-based network gave more dispersed results, but it could be noted that the IL6 interactome linked with Bilirubin degradation products *via* complement components, while Sphingomyelins and fatty acids formed separate interactomes **(Figure 4C)**.

**Figure 4:**
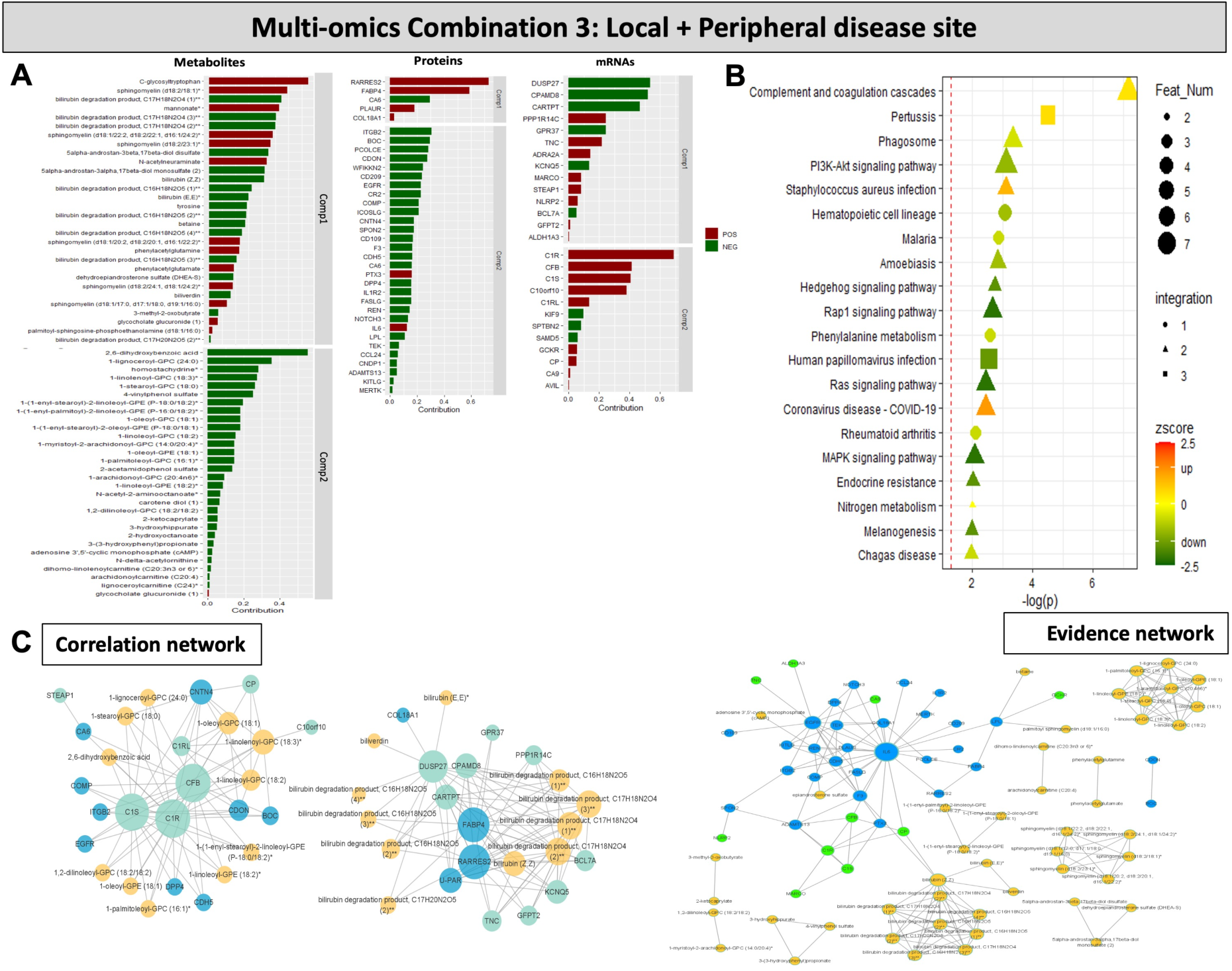
Multi-omics data integration combining local and peripheral disease site (Combination 3). **A)** Contribution of various metabolites, proteins and mRNAs from relevant single-omics layers to the integrated multi-omics model with 2 components. Bars represent the contribution of each feature to the symptomatic patient phenotype. Direction of contribution indicated with color (red-positive, green-negative). **B)** Joint enrichment analysis of all 3 omics layers in the multi-omics model. Circle indicates that the enriched term contains only one type of omics data, triangle represents terms containing 2 omics types, and square for 3 types. Direction of effect indicated with colour (green-to-red, legend). **C)** Correlation based network (rho >0.5) (to the left) and evidence-based network (to the right) of different analytical features from the multi-omics analyses. Color represents the type of omics data (blue, green and yellow referring to proteins, transcripts and metabolites respectively), size of the node represents the degree of significance.

#### Functional comparison of all three integrated multi-omics combinations points to ICOSLG and Sphingomyelins as common factors in local and peripheral disease sites

An overlap of the 3 integration combinations stratified by symptomatology, contained 6 commonly discovered analytes across all multi-omics combinatorial analyses. These were ICOSLG, several Sphingomyelins and 1-stearoyl-GPC **(Figure 5A)**. A detailed list of analytes from various multi-omics integrations is shown in **Supplementary Table VIII**.

**Figure 5:**
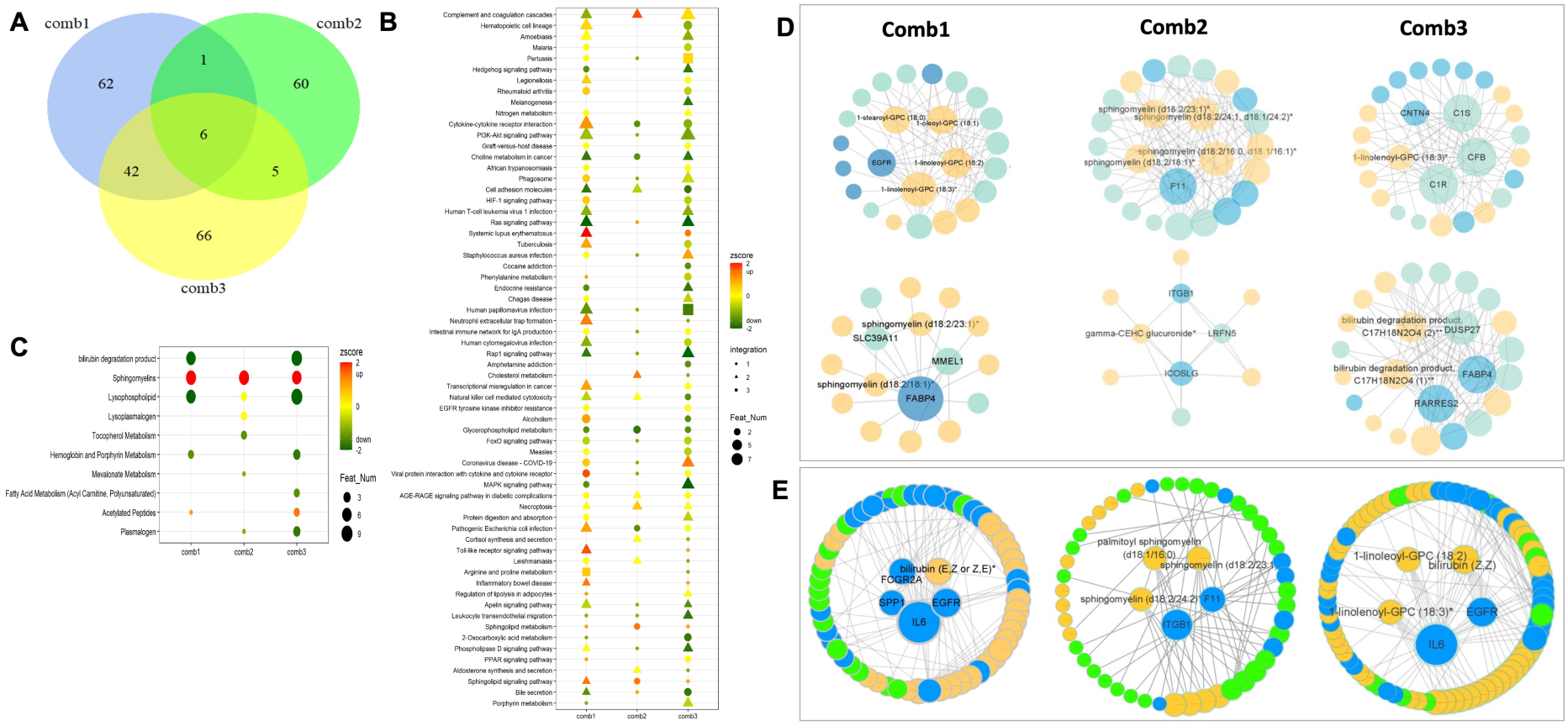
Functional comparison of all three integrated multi-omics combinations. **A)** Venn diagram showing numbers of analytes identified in each multi-omics combination, including a list of overlapping analytes. **B)** Joint and **C)** metabolomic data enrichment analyses for all 3 combinations, illustrating the number of different integrations (triangle, square, circle), direction of effect (color green-to-red) and the number of features per pathway (size of the node). Correlation **D)** or evidence-based **E)** functional network analyses of the 3 multi-omics integration combinations. Metabolites in yellow, proteins in blue, transcripts in green.

Joint gene set enrichment analysis revealed the importance of coagulation and necroptosis across all 3 combinations, but also activation of various immune processes and cholesterol metabolism as the most important pathways **(Figure 5B)**. Joint metabolomic analysis specifically confirmed the enrichment of Sphingomyelins and the repression of Bilirubin degradation products across all 3 integrations **(Figure 5C)**. Correlation- and evidence-based functional network analyses again highlighted the overall interplay among Sphingomyelins, coagulation factors and inflammation **(Figure 5D, E)**. In particular, network in Combination 1 (only peripheral plasma) was centered around IL6, EGFR, Bilirubin, SPP1, FABP4 and Sphingomyelins; network in Combination 2 (only local disease site) was centered around Sphingomyelins, ITGB1 and F11; while network in Combination 3 (the integration of plaque with peripheral plasma) centered again around IL6, EGFR and Bilirubin.

### Clinical, genetic and long-term follow-up studies for selected targets of interest

#### Analysis of selected targets with respect to cellular and morphological plaque features

We next interrogated the individual targets, selected from all 3 multi-omics integrations as the most significant and/or prominent in the evidence-based networks. BLVRA and BLVRB were selected as the key enzymes representing Bilirubin metabolism, which was highlighted in analyses of peripheral plasma along with FABP4 (Combination 1). ANGPTL3 was the gene that emerged from the local disease site studies (Combination 2), while ICOSLG and SMPD1 (a key enzyme in the Sphingomyelin metabolism) were selected as main genes from the intersection between local and peripheral disease sites (Combination 3). An *in-depth* analysis was performed for each of these top targets with respect to their expression levels across all datasets, association with plaque cells and morphological features, genetics, patient clinical chemistry parameters, as well as future outcomes, to assess their translational potential.

Significant upregulation of BLVRA and BLVRB transcripts was found in comparison of plaques from S *vs.* AS patients, and BLVRA was also dysregulated in PBMCs **(Figure 6A)**. Although plasma protein levels of these enzymes were not increased in this study **(Figure 6B)**, various components of the Bilirubin metabolism were detected in metabolomic analysis and all consistently showed downregulation in S *vs.* AS patients, both in local and in peripheral plasma **(representative metabolites in Figure 6C).** FABP4 transcript was significantly upregulated in plaques from S *vs*. AS patients, consistent with FABP4 protein being significantly increased in both local and peripheral plasma from S individuals. A significant downregulation of ICOSLG transcript and protein levels was detected in PBMCs, local and peripheral plasma from S *vs.* AS patients. While ANGPTL3 transcript was below the detection levels in plaques or in PBMCs, protein levels were significantly increased in peripheral and local plasma from S *vs.* AS individuals. Finally, SMPD1 transcript was detected at low levels and downregulated in PBMCs from S *vs*. AS patients. Protein levels of this enzyme were not analyzed in plasma in this study, but various components of the Sphingomyelin metabolism were all consistently upregulated in both local and peripheral plasma from S patients **(representative metabolites in Figure 6C).**

**Figure 6:**
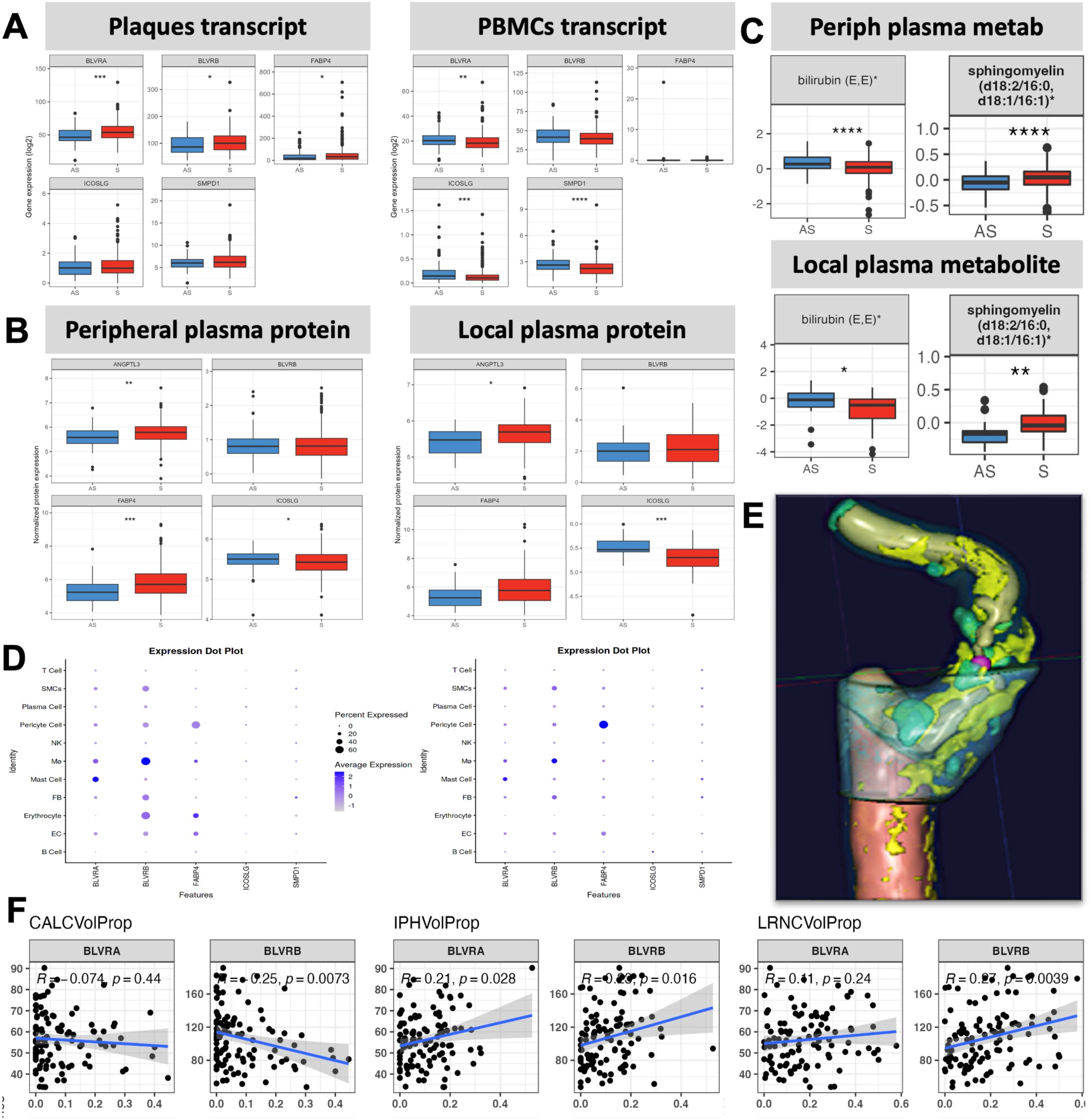
Expression analysis of selected targets of interest and association with plaque cellular composition and morphological features. **A)** mRNA expression of selected targets of interest BLVRA, BLVRB, FABP4, ICOSLG and SMPD1 in plaque and PBMC transcriptomic RNAseq data comparing S *vs.* AS patients. ANGPTL3 expression was not detected. **B)** Proteomic analysis of peripheral and local plasma comparing normalized levels of targets of interest in S *vs.* AS patients. **C)** Analysis of metabolomic data from peripheral and local plasma comparing normalized levels of representative bilirubin and sphingomyelin metabolism targets in S *vs.* AS patients. **D)** Carotid plaque scRNAseq data from *Alsaigh et al 2020* (left) and *Wirka et al 2019* (right) queried *via* the PlaqView open resource for cellular expression of selected targets. **E)** An illustration of the computed tomography (CT) carotid plaque image analyzed with the ElucidVivo software. Colors indicate different features, i.e. lipids are yellow, calcification turquoise, matrix is blue, etc. **F)** Correlation analyses were performed between the plaque RNA expression of selected targets and various quantified morphological features from diagnostic CTs (n=160 patients). IPHVolProp-intraplaque hemorrhage volume proportion, CalcVolProp-calcification volume proportion, LRNCVolProp-lipid rich necrotic core volume proportion.

Additionally, utilizing public scRNAseq datasets from human carotid plaques^31, 32^, we could further establish that BLVRA and BLVRB were both expressed in macrophages, as previously reported by us and others^11^, but the expression of BLVRB was considerably higher and more broadly associated also with erythrocytes, fibroblasts, pericytes and SMCs in plaques, while BLVRA was found also in mast cells **(Figure 6D)**. FABP4 seemed to be mostly expressed by pericytes, which has not been investigated so far, and lower levels were detected in macrophages, erythrocytes and endothelial cells. ICOSLG and SMPD1 were again expressed at low levels and could not be definitively localized in plaque cells, while ANGPTL3 was confirmed as undetectable.

The association of selected targets with plaque morphology features was next assessed by correlating BiKE RNAseq data with quantification of plaque tissue components from image analyses of pre-operative CTAs, as previously described **(Figure 6E and F)**^33–35^. Here, BLVRA and BLVRB significantly positively associated with total intraplaque hemorrhage volume proportion. BLVRB showed a positive association also with lipid rich necrotic core and a negative association with plaque calcification volume proportion. The other genes of interest did not show associations with plaque morphology in this analysis.

#### Association of selected targets with patient clinical biochemistry parameters

A detailed correlation analysis was performed between various blood parameters and transcript, protein or metabolite levels for selected targets **(Supplementary Figure VIII)**. Here, the most consistent correlations in plaques and plasma were found for BLVRB, FABP4 and ICOSLG. Our analysis suggested a significantly positive association for measures of kidney function with FABP4 and ICOSLG. Plasma levels of FABP4 were also positively associated with inflammatory parameters such as CRP, leucocyte count and fibrinogen. Another positive association was found between Hb1ac levels, and levels of BLVRB, FABP4, ICOSLG. Moreover, there was a significantly positive association between plasma levels of ANGPTL3 and total serum cholesterol in patients.

With respect to metabolites, positive association was found between Bilirubin levels and Bilirubin degradation products, as expected **(Supplementary Figure VIII)**. These metabolites were also consistently positively correlated to Hb levels and erythrocytes, but negatively to CRP, HbA1c and fibrinogen, and without correlations to lipid parameters. On the other hand, plasma levels of Sphingomyelins correlated positively with most lipid parameters (total cholesterol, LDL, HDL, except TG) and adiponectin levels, but negatively with Hb and HbA1C. These correlations suggest possible mechanistic dependencies among the targets of interest and classical, clinically used blood markers of various cells and processes.

#### Association of selected targets with medications

Information about relevant medications, i.e. lipid-lowering, anti-diabetic, anti-hypertensive and anti-thrombotic therapies, was available in BiKE for the majority of patients and was tested in association with the levels of various analytes in plaques, PBMCs and plasma (selected data shown in **Supplementary Figure IX)**. After adjustment for age and sex, significant results could be shown for the transcript expression of SMPD1, ICOSLG and BLVRB in peripheral PBMCs and treatment with anti-thrombotics, anti-hypertensives and in particular lipid-decreasers, where the association with ICOLSG was strongly significant. On protein level, anti-hypertensive medication showed an effect on FABP4 in both local and peripheral plasma. On metabolite level, anti-hypertensive medication also showed marginal association with plasma Bilirubin, while anti-diabetics had an impact on plasma Sphingomyelins. Of interest, plasma ANGPTL3 protein levels were not affected by any of the tested medications.

#### Association of selected targets with long-term survival of patients

Survival has been monitored for >15 years in BiKE patients^30^, with respect to the cumulative MACCEs (MI and ischemic stroke events), as well as all cause death. This allowed us to interrogate the association between levels of selected targets and adverse outcomes, which could suggest biomarkers of long-term risk in patients already treated with CEA.

Analysis showed that patients with higher peripheral plasma protein levels of ANGPTL3 (top quartile) suffered from significantly more MACCEs and all cause death, showing an overall vulnerable profile **(Figure 7)**. Lower (bottom quartile) Bilirubin metabolite peripheral plasma levels were indicative of MIs and all cause death **(Figure 7)**, but lower plaque BLVRB expression was associated with less MACCEs and MIs **(Supplementary Figure X)**. Of note, BLVRA did not show any significance in this analysis. Lower levels of various Sphingomyelins in peripheral plasma systematically associated with more MACCEs and ischemic strokes **(Figure 7)**. Higher expression of ICOSLG in plaques was linked with poorer survival with respect to MACCEs and ischemic stroke, but FABP4 expression did not show any significance in these analyses **(Supplementary Figure X)**.

**Figure 7:**
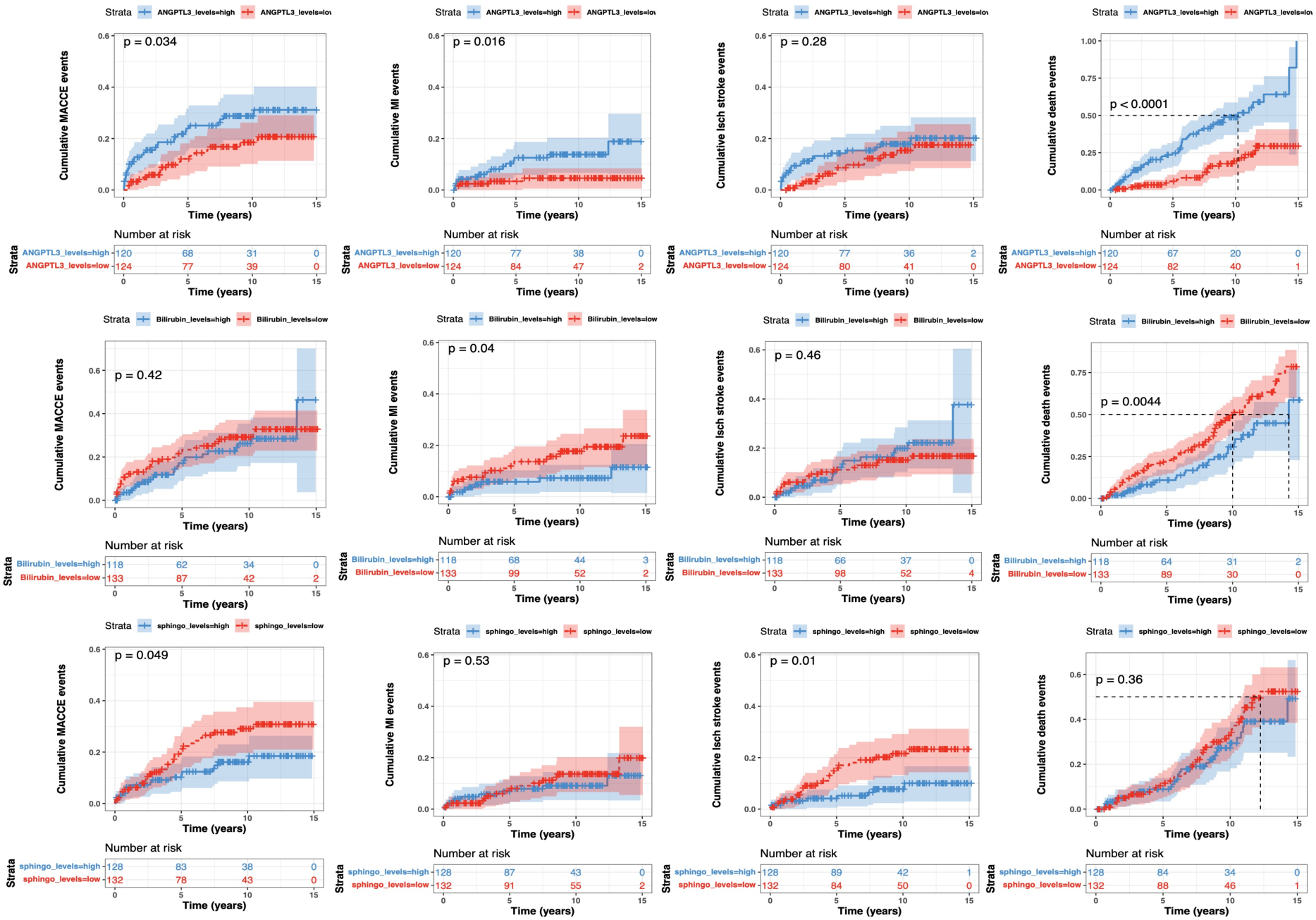
Cumulative Kaplan-Meier outcome estimates after carotid surgery. Plots illustrating survival of CEA patients during the 15 years follow-up period after surgery, based on top *vs.* bottom quartile of relevant analyte levels in peripheral plasma. Both patients that were symptomatic and asymptomatic at surgery were included in the follow-up analysis. Each mark along the lines indicates an event, numbers at risk indicated in tables under the plots. Top raw shows plots for peripheral plasma ANGPTL3 protein levels. Middle raw shows plots for Bilirubin metabolite peripheral plasma levels, and bottom raw Sphingomyelin metabolite peripheral plasma levels. MACCEs-major adverse cardio- and cerebro-vascular events; MI- myocardial infarction.

#### Prioritized targets show genetic linkage with cardiovascular disease traits

Finally, previously prioritized candidates (FABP4, BLVRA, BLVRB, SMPD1, ICOSLG, ANGPTL3) were extended with other top analytes from each of the multi-omics integrations (F11, IL6, ITGB1, MPZL1, SOX11, TFP, C1R, C1S, CFB, EGFR, SPP1), together forming a list of 17 candidates selected for genetic association analyses. We tested for target-CVD associations using the public OpenTargets platform, a powerful tool for drug discovery that, by integrating diverse data sources like genomics, proteomics, literature, chemical databases and clinical trials, can identify genes potentially involved in cardiovascular conditions. It was noteworthy that 5 out of these 17 targets showed the overall global association score >0.4 for CVD (on a scale of 0-1, with 1 being the strongest association), with F11, C1S, EGFR, IL6, ANGPTL3 being the top on this list. Additionally, 4 out of these top targets (F11, C1S, EGFR and IL6) showed strong and likely causal contribution of genetics in CVD **(Supplementary Table X)**. Of note, we tested for traits like atherosclerosis, carotid atherosclerosis and coronary artery disease in a similar way, but such specific analysis did not reveal strong target-disease associations.

## Discussion

Single- and multi-omics analyses were performed on data from a large biobank of carotid atherosclerosis, with focus on instability as defined by patient symptomatology. Key pathways and molecular signatures of unstable disease were confirmed, such as coagulation, necroptosis, immune activation, cholesterol metabolism, while less explored ones were also highlighted, i.e. heme- and Bilirubin degradation and Sphingomyelin metabolism. In peripheral plasma, FABP4, IL6, Bilirubin and Sphingomyelin metabolism were the most prominently enriched analytes, while F11, ANGPTL3, ICOSLG, ITGB1, Sphingomyelin were enriched in the local disease site. FABP4, C1R, IL6, Bilirubin and Sphingomyelin also appeared at the intersection of both disease locations. Extended clinical analyses showed an impact of the lipid-lowering therapy on ICOSLG levels in PBMCs, anti-hypertensives on plasma FABP4 and BLVRB, while anti-diabetics had an impact on plasma Sphingomyelins, however none of the medications seemed to affect plasma ANGPTL3 levels. Moreover, association with future adverse events or death could be shown for peripheral plasma levels of ANGPTL3, Bilirubin and Sphingomyelin, as well as for ICOSLG plaque transcript levels. Target-disease analyses revealed likely causal genetic links for F11, C1S, EGFR and IL6 with CVD.

The single-omics analyses performed in this study using state-of-the-art technologies, are some of the most comprehensive ever reported in the field, illuminating key molecular features of unstable atherosclerosis. Plaque transcriptomics confirmed important pathways in plaque instability such as epithelial-to-mesenchymal transition, TNFa and IL6 signalling, IFNg response, hypoxia and angiogenesis, ECM reorganisation and bone remodelling, cholesterol homeostasis and glycolysis. Many of these pathways were also found in PBMC transcriptomics, in addition to coagulation, heme-metabolism and angiogenesis that seemed more specifically associated with circulating cells. Plasma proteomics confirmed the overall upregulation of chemokine signalling and cytokine-receptor interactions, while IL17 pathway was particularly enriched in local plasma retrieved from the vicinity of plaque at surgery. Plasma metabolomics revealed an increase in Sphingomyelins in both local and peripheral samples, while Bilirubin metabolites were significantly repressed in peripheral plasma. The metabolomics dataset is novel and potentially of high relevance for future studies in the field, considering that atherosclerosis is linked with systemic metabolic dysregulation^36^. The rapid growth in metabolomics platforms has only just begun to provide insights into the new pathophysiological pathways for CAD, cardiomyopathies, stroke, and diabetes^37–41^. Moreover, the power of pharmacometabolomic research has recently been demonstrated, when it was reported that baseline plasma levels of a single metabolite 2-hydroxyvaleric acid, can discriminate between responders and non-responders to statins^41^. Nevertheless, single-omics technologies remain insufficient in capturing the complex intricacies of human disease biology.

In recent years, multi-omics has shown promise for providing more robust subsystem classifiers and help in elucidating interactome interplay around the phenotype of interest^28^. The complex, multi-factorial nature of CVD makes it difficult to recapitulate the key molecular drivers and signalling pathways without system level approaches. This notion is strongly emphasised in our study, as both well-known and novel molecular targets and mechanisms appeared from each of the three different multi-omics integration combinations. Integration of data in Combination 1 from peripheral circulation solely was intended to yield potential biomarker discoveries. Indeed, IL6 as one of the top targets that emerged from this integration, is widely supported by numerous previous reports associating it with adverse cardiovascular outcomes, both in subjects with and without a history of CVD^42^. Combination 2 integrated only data from the local disease site that we have previously shown to be enriched with lesion-derived analytes^11^. Here, candidates such as F11 and ANGPTL3 appeared, possibly reflecting their association with coagulation/hemorrhage, and lipid accumulation in plasma in the vicinity of the lesion, respectively^43, 44^. Finally, Combination 3, integrating plaque with peripheral circulation data aimed to enrich our results for blood signatures specifically reflecting carotid plaque phenotype. This analysis showed that there was at least a portion of molecular signatures related to the lipid metabolism (i.e. FABP4, Sphingomyelins), coagulation (C1R), inflammation (IL6), but also signatures of hemoglobin degradation (Bilirubin), which were detectable in peripheral plasma and possibly reflect instability processes associated with intraplaque hemorrhage. Together, these well-known and novel markers constitute prioritized candidates for future development of biomarkers and pharmacotherapy.

Here, we initiated their validation by performing translational analyses in patient clinical data. Our results showed that various cell types could be the sources of FABP4 in both plaques and plasma. FABP4 increase in plasma could potentially reflect systemic inflammation, diabetes and kidney dysfunction, as well as hypertension, indicated by the finding that anti-hypertensive medication showed an effect on its plasma protein levels. From these observations, it is possible that measurement of FABP4 levels could be used to improve patient phenotyping. Previous studies have already reported increased expression of FABP4 in carotid plaques, association to adverse events on short-term follow up, and suggested that it could be a key factor connecting vascular lipid accumulation with inflammation^45, 46^.

ICOSLG is a ligand for the T-cell-specific surface receptor ICOS, whose expression is enhanced by TNF. It acts as a costimulatory signal for T-cell proliferation and cytokine secretion and serves an important role in mediating local tissue responses to inflammatory conditions^47^. In our study, ICOSLG was generally repressed in plaques, PBMCs and plasma from S patients on cohort level, but in those patients with a relatively higher expression in their plaques, it was associated with future adverse events. In PBMCs, its expression was strongly affected by lipid-lowering medication. It has previously been shown that blocking the ICOS/ICOSLG pathway with ICOSLG antibodies reduced the development of atherosclerosis coupled to the formation of tertiary lymphoid organs^47^. ICOS/ICOSLG pathway has been suggested as a promising target for a new wave of immunotherapies, confirmed in our studies.

ANGPTL3 is a target that was solely identified in plasma, enriched in association with S disease and blood levels of all major lipid fractions, where high levels of ANGPTL3 were a strong predictor for poor long-term survival. ANGPTL3 is a factor secreted by the liver that inhibits LPL and other lipases *via* the formation of a complex with related protein ANGPTL8^48^. Genetic studies have shown that carriers of loss-of-function variants in ANGPTL3 have lower plasma LDL and triglyceride levels, and are at reduced risk of ASCVD^48^. Clinical studies in patients with different forms of dyslipidemia have demonstrated that inactivation of ANGPTL3 using monoclonal antibodies (evinacumab) or antisense oligonucleotides markedly lowers plasma LDL and TG levels^49^. Our study adds to the confirmation of these results specifically in the context of carotid atherosclerosis and suggests that the beneficial effects of clinical ANGPTL3 inhibition should be evaluated broadly across the different CVD pathologies. Even more so, as we could not show any effect of currently available medications on plasma ANGPTL3 levels.

Bilirubin is the ubiquitous end-product of heme catabolism, necessary for clearance of waste products from turnover of red blood cells^11^. We have previously shown that Biliverdin Reductase B (BLVRB), an enzyme converting Biliverdin to Bilirubin, is a novel marker of intraplaque haemorrhage and a predictor of ischemic stroke in patients with symptomatic carotid atherosclerosis^11, 50^. In the present study, Bilirubin metabolites, as well as BLVRA and BLVRB, were strongly dysregulated in various omics datasets, however in opposite trends. Both enzymes were enriched in plaques from S patients, with macrophages as the likely source, and showed association with intraplaque hemorrhage on preoperative CTA scans. On the other hand, metabolites associated with this pathway were all repressed in both local and peripheral plasma, in S *vs.* AS patients, and overall positively associated with Hb and various erythrocyte parameters. Furthermore, anti-hypertensive medication showed a marginal effect on plasma Bilirubin levels in BiKE patients. Despite the yet unclear mechanistic relationships with CVD, the importance of this pathway has been suggested also from Mendelian randomization studies in the UK biobank, where genetically determined plasma Bilirubin levels have been negatively associated with the risk of CVD^51^. Together, this suggests a complex feedback relationship between enzymes related to Bilirubin generation in plaques and Bilirubin metabolites detected in plasma, which originate mostly from liver. However, this relationship could also be linked to intraplaque haemorrhage as a key feature of unstable atherosclerosis^50^, especially since we observed that levels of Bilirubin metabolites in peripheral plasma were indicative of MIs and all cause death.

Sphingomyelin metabolites were strongly enriched in both peripheral and local plasma in S patients, where they correlated positively with various lipid parameters, but also seemed to be affected by anti-diabetic medication. However, it may be that the role of Sphingomyelins is protective in this context, as their lower plasma levels systematically associated with more MACCEs and IS. Sphingomyelins are one of the classes of sphingolipids that have been described as elevated in S *vs.* AS patients^52, 53^. The key enzyme involved in sphingomyelin hydrolysis is acid sphingomyelinase (ASM), coded by the gene *SMPD1*^54^, which when deficient, causes Neimann-Pick disease, characterized by toxic Sphingomyelin and lipid accumulation in various organs, linked to an atherogenic lipid profile and increased risk for atherosclerosis ^55, 56^. This clinical observation together with the strong association between sphingolipids and CVD risk, has motivated the development of a sphingolipid-inclusive CAD score, which has been suggested to be superior to conventional clinical biomarkers^57^. Our results support this by confirming the association of Sphingomyelins with atherosclerosis and extending their prognostic value in this context.

Moreover, genetic evidence has been observed in our study for targets such as IL6 and F11 in association with CVD. Circulating levels of IL6 have been widely associated with different CVDs and outcomes, i.e. with the risk of atrial fibrillation. But also, IL6 signaling has been associated in a causal manner with the risk of stroke independently from atrial fibrilation^58^, therefore it is not surprising that IL6 came up as one of the top candidates in our carotid cohort stratified for cerebrovascular events. Previously, genetic variants in F11 have been associated with venous thromboembolism on GWAS level^59^, but recently have also been suggested as a prognostic factor in ischemic stroke of arterial origin^60^. Although historically viewed as distinct disorders with respect to etiology, pathophysiology and treatment, evidence is emerging that thromboembolic events in veins and arteries share some genetic and mechanistic features. Possibly, F11 could be at the center-point of these commonalities, as our study suggests also its genetic linkage with CVD.

Finally, the inclusion of metabolomics data contributed towards novel discoveries in the network analyses of our multi-omics integrations, where entirely novel molecular interactomes emerged. For example, in peripheral circulation we found a direct and previously never explored association of FABP4 with various Sphingomyelins and Bilirubin, while IL6 showed links with Bilirubin degradation products. In the local disease site, there was an association of F11 and ANGPTL3 with various Sphingomyelins and cholesterol. At the intersection of plaque and peripheral disease site, a novel network appeared where several complement factors (C1S, C1R, CFB) associated with various fatty acids. Here again, IL6 interactome was linked with Bilirubin degradation products *via* complement factors, while Sphingomyelins and fatty acids formed separate interactomes. These novel networks and previously unexplored interactomes are mechanistically interesting for further studies because they could harbor significant potential for identifying biomarker-drug combinations to monitor unstable clinical atherosclerosis phenotypes.

### Limitations

The choice of software and parameters for multi-omics integration could have affected the results, although DIABLO, which is a supervised approach, has been widely benchmarked for these types of analyses. In multi-omics integration, unsupervised methods are also often used, however, given the rich nature of the omics and clinical data available in BiKE, such choice could have led to even more complex information. Hence, we opted for a reductionist approach with DIABLO, which has been used for similar biobank studies earlier^61^.

Another general challenge in integration across multiple platforms is the issue of data missingness, since technical limitations result in different omics datasets not being able to identify the corresponding analytes across all layers, essentially confounding the analysis towards the smallest, most restrictive omics dataset. Rationalizing this discordance among different omics data, whether it may be attributable to sample size, patient subphenotypes, technical variability, or differential regulation of gene *vs*. protein *vs*. metabolite networks, is key to the analysis. Interpreting the directionality and causality of relationships within multi-omic data is another important challenge, where our results will require experimental validations. However, it is worth noting that some of these limitations are mitigated by the large size of BiKE datasets, as well as the confirmation of many targets already in clinical trials.

Even though BiKE single- and multi-omics analysis, clinical associations and survival analysis from this cohort, in combination with OpenTarget platform genetic linkage, jointly provided a systematic triaging for prioritized CVD targets, our cohort size is still limited to infer strong causal claims due to the lack of its own genetic data layer.

Finally, the BiKE biobank exclusively contains end-stage atherosclerosis patients, with a relatively narrow age-span, which constricts our findings and limits extrapolation of conclusions into processes relevant for earlier stages of atheroprogression. Although BiKE cohort is large and well-phenotyped, it is also rather homogeneous in its composition and dominated by Caucasian descent. Therefore, results from this study should be validated in larger atherosclerosis cohorts with diverse ethnic backgrounds, where stringent corrections for multiple comorbidities can be performed.

### Conclusions and outlook

In summary, this study generated some of the largest, state-of-the-art omics datasets in the field and performed first-of-a-kind multi-omics integration in human carotid atherosclerosis. Our results validate numerous previously reported pathways enriched in atherosclerotic plaques and plasma, but also reveal novel targets and pathways from the integration between local vascular disease site and peripheral circulation. The endeavors presented here could have paradigm-shifting implications, especially for enabling tailored therapies and precision medicine in the fight against carotid atherosclerosis, including transfer of generated knowledge to coronary and lower limb territories **(Visual Abstract)**. As statins remain a cornerstone of treatment for all these patients, new generation of therapeutic modalities complementing or enhancing statin effectiveness and companion biomarkers, could revolutionize CVD management. By applying multi-omics to decipher the intricate systems architecture of atherosclerosis, our study provides a platform for forging novel biomarker-drug combinations that can reflect the state of plaque composition or vulnerability, and herald a new era of personalized cardiovascular care, ensuring better outcomes for patients worldwide.

## Supporting information

Multi-omics Supplementary File BiKE

Multi-omics Supplementary Tables BiKE

## Acknowledgements

DNA and RNA sequencing were performed by the SNP&SEQ Technology Platform in Uppsala, Sweden. The facility is part of the National Genomics Infrastructure (NGI) supported by the Swedish Research Council for Infrastructures and Science for Life Laboratory, Sweden. The authors would also like to acknowledge support of the National Genomics Infrastructure (NGI)/Uppsala Genome Center and UPPMAX for aiding in computational infrastructure and basic data QC analyses. The SNP&SEQ Platform is supported by the Swedish Research Council and the Knut and Alice Wallenberg Foundation, while NGI/Uppsala Genome Center has been funded by the Swedish Research Council and Science for Life Laboratory, Sweden.

## Disclosure of interest

VD, jDj, NBM and KCK are employees of Novo Nordisk A/S Måløv, Denmark. They hold minor stock portions as part of an employee offering program. The other Authors have nothing to declare.

## Funding

This study was performed as part of the strategic research partnership between Ljubica Matic/Ulf Hedin, KI, Sweden and Novo Nordisk, Denmark, under the agreement number DNR 4-3642/2019 and data transfer agreement number DNR 4-1141/2021. Ljubica Matic is the recipient of fellowships and awards from the Swedish Research Council [VR, 2023-02724, 2019-02027], Karolinska Institute Consolidator program, Swedish Heart-Lung Foundation [HLF, 20240094, 20241083, 20230357, 20210466, 20200621, 20200520, 20180244, 20180247, 201602877], Swedish Society for Medical Research [SSMF, P13-0171]. Ljubica Matic also acknowledges funding from Mats Kleberg’s, Sven and Ebba-Christina Hagberg’s, Tore Nilsson’s, Magnus Bergvall’s and Karolinska Institute research (KI Fonder) and doctoral education (KID) foundations. Project funding was also obtained by Ulf Hedin from the Swedish Heart-Lung Foundation (20180036, 20170584), the Swedish Research Council (2017-01070, 2019-02027), and King Gustav Vth and Queen Victoria’s Foundation. This work was also funded by a research grant from the European Union’s HORIZON-HLTH-2023-TOOL-05 program with Grant agreement No 101136962 (NextGen).

## Author contributions

LM and NBM have set up all BiKE omics data profiling (transcriptomics, proteomics and metabolomics) *via* core facilities and external vendors. MK performed biobank material processing and preparation for analyses. VD, ZX, DjDj, OB, AJB, SN and LM performed data collection, pre-processing and/or statistical and bioinformatic analyses. VD, DjDj, NBM, KCK, UH and LM conceived the study, provided expert guidance, methodological and/or conceptual input. VD and LM designed and supervised the study and interpreted results. LM drafted the first version of the manuscript. All authors provided comments and participated in critical revisions of the manuscript.

## Data availability statement

All data and results generated or analyzed during this study are included in this article and/or the Supplementary Materials. Created codes are publicly available at: https://github.com/vd4mmind/BIKE_Athero. The individual human data reported in this study cannot be deposited or shared because of the GDPR and ethics laws that regulate the privacy of individuals that participated in the BiKE study. Other data reported in this paper on group level, as well as resources and reagents, may be shared upon a reasonable request to the Corresponding Author.

## Notes

### Competing Interest Statement

VD, DjDj, NBM and KCK are employees of Novo Nordisk A/S Malov, Denmark. They hold minor
stock portions as part of an employee offering program. The other Authors have nothing to
declare.

### Funding Statement

The study was funded by multiple organizations as listed below
Novo Nordisk A/S Denmark
Swedish Research Council and Karolinska Institute Consolidator program Swedish Heart-Lung Foundation
Swedish Society for Medical Research
Karolinska Institute research (KI Fonder) and doctoral education (KID) foundations.
Swedish Heart-Lung Foundation
Swedish Research Council
King Gustav Vth and Queen Victoria Foundation
European Union HORIZON-HLTH-2023-TOOL-05 program

### Author Declarations

Studies within BiKE were approved by the Regional Ethical Committee of Stockholm (DNRs 95-277, 95-276, 01-199, 02-146, 02-147, 2017/505-32) and conducted in accordance with guidelines of the Declaration of Helsinki. All human samples and personal data were collected with informed consent from patients or organ donors guardians. The reporting of this study is compliant with the STROBE guidelines.

