## Supplementary material for "Multi-omics data integration from patients with carotid stenosis illuminates key molecular signatures of atherosclerotic instability": Multi-omics Supplementary File BiKE

### Supplementary Material and Methods

##### Human carotid plaque cohort (Biobank of Karolinska Endarterectomies)

Patients undergoing carotid endarterectomy (CEA) at the Departments of Vascular Surgery from both Karolinska University Hospital or Södersjukhuset (affiliated to BiKE in 2008), Stockholm, Sweden were consecutively included in the Biobank of Karolinska Endarterectomies (BiKE) and clinical data recorded upon admission. Symptomatic (S) patients were categorized as those having a minor stroke (MS), transient ischemic attack (TIA), or ipsilateral ocular symptoms (*amaurosis fugax*, AFX), while patients without qualifying symptoms within 6 months prior to surgery were considered asymptomatic (AS). Plaques and blood were collected during surgery and retained within BiKE. "Local" samples were retrieved by aspirating blood from the carotid artery lumen adjacent to the plaque 5 minutes after artery cross clamping and before arteriotomy, as previously described.[12] Plaques were divided transversally at the most stenotic part and the proximal half of the plaque was frozen at -80°C immediately after surgery and later used for transcriptomic or proteomic profiling. The distal half was fixed in 4% zinc-formaldehyde and processed for histology. Blood was processed according to standard procedures for separation of monocyte cell fraction and RNA isolation. Briefly, peripheral blood mononuclear cells (PBMCs) were isolated from blood collected at surgery *via* density gradient centrifugation through Ficoll-Paque (Vacutainer CPT, Becton-Dickinson, Franklin Lakes, NJ). PBMCs were processed using RLT buffer (Qiagen, Valencia, CA) before freezing at -80°C. BiKE is today one of the largest tissue cohorts for carotid disease and has been extensively described in numerous prior publications[1].

##### BiKE clinical, epidemiological and follow-up data collection

Individual data for death, major adverse cardio- and cerebro-vascular events (MACCEs), comorbidities and blood parameters were extracted from the national registries (i.e. Swedish Cause of Death Register, Swedish Hospital Discharge Register, etc.), clinical patient charts and BiKE database and stored using a system of individual pseudonymization for compliance with the General Data Protection Regulations (GDPR). Individual data such as diagnostic codes from the International Classification of Diseases (ICD-10), were extracted for every hospitalization and visit to the specialist outpatient clinic from the National Patient Register. The following data were extracted from the clinical database of BiKE and medical records: patient's smoking habits, symptomatology prior to surgery, body mass index, hypertension, kidney function and medications. The start of inclusion was 31$^{st}$ August 1998, and the end was 12$^{th}$ December 2017. Data were extracted from national registries on 22$^{nd}$ January 2018. Follow-up was calculated from the date of the patient's first carotid surgery. Patients treated with CEA bilaterally were included at the first procedure.

##### Ethics statement

Studies within BiKE were approved by the Regional Ethical Committee of Stockholm (DNRs 95-277, 95-276, 01-199, 02-146, 02-147, 2017/505-32) and conducted in accordance with guidelines of the Declaration of Helsinki. All human samples and personal data were collected with informed consent from patients or organ donors' guardians. The reporting of this study is compliant with the STROBE guidelines.

#### Transcriptomic analyses - *RNA sequencing*

RNA was prepared using Qiazol Lysis Reagent (Qiagen, Hilden, Germany) and purified by RNeasy Mini kit (#74106, Qiagen, Germany), including DNase digestion. The concentration was measured using Nanodrop ND-1000 (Thermo Scientific, Waltham, MA) and quality estimated by a Bioanalyzer capillary electrophoresis system (Agilent Technologies, Santa Clara, CA). The library for bulk RNA sequencing of RNA from plaques and PBMCs was prepared using TruSeq stranded total RNA with RiboZero Globin treatment (# 20020612/20020613, Illumina Inc.). Paired-end 150bp read length, NovaSeq 6000 system, S4 flow cells and v1 sequencing chemistry was used for sequencing at 20 mreads/sample. Libraries that yielded less data then aimed for were re-sequenced on SP flowcell. The Bcl to FastQ conversion was performed using bcl2fastq_v2.20.0.422 from the CASAVA software suite, followed by downstream analysis. The raw fastq files were processed using nf-core RNAseq pipeline. Human reference genome GRCh38 obtained from Ensembl was used for alignment. Gene level counts data of protein-coding and lincRNA genes were considered for all the downstream analysis. R package DESeq2 v1.34.0(51) was employed for differential expression analysis with adjustment for confounding factors such as age and gender performed using R/Bioconductor package DESeq2 v1.26.0. Pathway enrichment analysis was performed using the tool GSEA v4.1.0.

#### Proteomic analyses - *Plasma proteomics*

The following five Olink® panels were used in plasma profiling: *Olink® Target 96 Cardiometabolic*, *Olink® Target 96 CVD II*, *Olink® Target 96 CVD III*, *Olink® Target 96 Development*, and *Olink® Target 96 Immuno-Oncology*. This multiplex biomarker platform detected approximately 500 proteins in plasma. The platform uses proprietary Proximity Extension Assay (PEA) technology and a readout based on Next Generation Sequencing (NGS) in Illumina NovaSeq 6000. OlinkAnalyze package was used to perform the standard quality control (QC) on this panel of protein analytes using the NPX function and relevant QC parameters as suggested by the package vignette to remove the outlier samples that do not meet their standard quality flags. For multivariate analysis to identify differential proteins between S *vs.* AS groups, we used generalized linear mixed model package lme4. This analysis was run across 2 different sites, peripheral (EP) and local (STP). The final models were adjusted for sex + age. Only proteins with p value ≤ 0.05 were chosen for subsequent downstream analysis.

#### Metabolomic analyses – *Plasma metabolomics*

Plasma metabolomic analyses were performed at Metabolon, Morrisville, NC. Samples were prepared using the automated MicroLab STAR® system from Hamilton Company. To remove protein, dissociate small molecules bound to protein or trapped in the precipitated protein matrix, and to recover chemically diverse metabolites, proteins were precipitated with methanol under vigorous shaking for 2 min (Glen Mills GenoGrinder 2000) followed by centrifugation. The resulting extract was divided into five fractions: two for analysis by two separate reverse phase (RP)/UPLC-MS/MS methods with positive ion mode electrospray ionization (ESI), one for analysis by RP/UPLC-MS/MS with negative ion mode ESI, one for analysis by HILIC/UPLC-MS/MS with negative ion mode ESI, and one sample was reserved for backup. Samples were placed briefly on a TurboVap® (Zymark) to remove the organic solvent. Experimental samples were randomized across the platform run with QC samples spaced evenly among the injections. All methods utilized a Waters ACQUITY ultra-performance liquid chromatography (UPLC) and a Thermo Scientific Q-Exactive high resolution/accurate mass spectrometer interfaced with a heated electrospray ionization (HESI-II) source and Orbitrap mass analyzer operated at 35,000 mass resolution.

Raw data was extracted, peak-identified and QC processed using Metabolon's hardware and software. Compounds were identified by comparison to library entries of purified standards or recurrent unknown entities. Metabolon maintains a library based on authenticated standards that contains the retention time/index (RI), mass to charge ratio (m/z), and chromatographic data (including MS/MS spectral data) on all molecules present in the library. Furthermore, biochemical identifications are based on three criteria: retention index within a narrow RI window of the proposed identification, accurate mass match to the library +/- 10 ppm, and the MS/MS forward and reverse scores between the experimental data and authentic standards. The MS/MS scores are based on a comparison of the ions present in the experimental spectrum to the ions present in the library spectrum.

The present dataset comprises a total of 1090 compounds of known identity (named biochemicals). Following log transformation, we again used lme4 as for proteomics, for multivariate analysis to identify differentially regulated metabolites between S *vs.* AS groups across EP and STP disease sites. The models were adjusted for sex and age. Only metabolites with p value ≤ 0.05 were chosen for subsequent downstream analysis with MetaboAnalyst 5.0 for functional enrichment.

#### Multi-omics data integration using DIABLO

DIABLO software from the mixOmics R package (version 6.25.1) was used for the entire analysis[2]. Normalized data across each omics type (transcriptomics, proteomics, metabolomics) were used as input for supervised DIABLO. The analysis was performed for 3 different combinations (peripheral circulation – Combination 1, local disease site – Combination 2, local plaque with peripheral circulation – Combination 3) to compare and identify cross-sectional molecular analytes that capture the difference between S *vs.* AS groups. Overall greater than 3000 features from across all omics types were used for analysis with DIABLO in each combination. Optimal number of supervised features across each omics type was determined by tune.block.spslda function. Greater than 28000 models have been fitted for each component and each nrepeat, using "centroid.dist" parameter with a 10-fold cross validation to identify the optimal number of features across transcripts, proteins and metabolites used for downstream integration analysis. A systematic assessment around the design matrix was performed that assumes the contribution from each omics layer for integration and feature selection between 0%, 10%, 20%, 30% and 40%. The final model was selected and fitted to block.splsda with a weighted design matrix of 30% contribution across omics types that can discriminate and explain the phenotypic variance. nComponents parameter used in this analysis was 3, with 10-fold cross-validation. While performing any downstream analysis around enrichment of identified analytes, only those that had correlation coefficient ≥ 0.5 were selected.

#### Target mining and online bioinformatic tools

A list of all significantly differentially regulated molecules/analytes (genes, transcripts, metabolites or proteins) after corrections for multiple comparisons, was assembled from single- and multi-omics analyses. This list contained originally n=268 analytes from single-omics and n=111 from multi-omics **(Supplementary Table IX).** The list was then enriched with information from public databases on e.g. tissue-wide expression and localisation (GTex, HPA), plaque scRNAseq cellular fractions (PlaqView), druggability and epigenetic information (Enrichr), PubMed literature mining related to atherosclerosis/CVD, or data from animal models of atherosclerosis/CVD. The StringDB database was used to build protein-protein interaction networks for targets of interest.

Selected targets were checked for gene/protein-disease association with CVD traits using OpenTargets platform[3]. The platform provides evidence and global association score on user-defined targets to diseases of interest like atherosclerosis, coronary artery disease, heart failure or cardiovascular diseases in general. These scores can be further prioritized based on the strength of evidence, for hypothesis generation and drug development efforts.

**Enrichment analyses and visualization**

Enrichment analysis was performed using clusterProfiler v4.8.2 based on databases (metabolon annotation list, KEGG, SMPDB, GO, Hallmark, Reactome pathways, WikiPathways). Joint enrichment analysis was performed using MetaboAnalyst 5.0; clusterProfiler v4.8.2 based on databases (KEGG, SMPDB). R package ggplot2 v3.3.2 was used to create bubble plots, scatter plots, and boxplots. R/Bioconductor package ComplexHeatmap v2.2.0 was used to create all heatmaps. Network analysis was performed by tools and databases (omicsnet 2.0, metaboanalyst 5.0, metabolon annotation list). Network diagrams were drawn in Cytoscape ver 3.8.2 and hubs identification by cytoHubba (Cytoscape Plugin). Venn diagrams were generated using the online tool InteractiVenn.

**Computed tomography angiography (CTA) and image analysis**

Carotid CTAs was performed pre-operatively as previously described[4, 5]. Reconstructed images (0.625 mm) were analysed by trained observers using the ElucidVIVO® (Elucid Bioimaging Inc., Boston, MA) software as previously described[4-6]. In brief, the external carotid artery was excluded from analyses and the lumen and wall of the common and internal carotid artery evaluated automatically where the software creates 3D segmentations with voxel-wise discrimination of intraplaque tissue types including lipid-rich necrotic core (LRNC), calcification (CALC), intraplaque haemorrhage (IPH), and extracellular matrix (MATX, representing plaque tissue not identified as any of the previous components). The proportion of these components relative to the total wall volume was quantified (VolProp) together with structural features such as wall volume plaque burden (proportion of total vessel volume or area occupied by the plaque), minimal fibrous cap thickness (shortest distance from the edge of the LRNC to the lumen in microm), and degree of stenosis (NASCET).

**Survival analyses**

The incidence for outcomes was plotted with Kaplan-Meier curves, with differences tested by log rank test. Survival curve was plotted using the number of myocardial infarctions (MI), ischemic strokes (IS), major cardiac and cerebral events (MACCE) and all-cause death at each unique time point for a period of 15 years after the patients have undergone carotid endarterectomy. To perform survival analysis, the patients were stratified according to the 25$^{th}$ quartile (low) and 75$^{th}$ quartile (high) of the relevant analyte levels.

#### Supplementary Tables and Table legends

All Supplementary Tables are available as a searchable Excel file.

#### Supplementary Figures and Figure legends

#### Supplementary Figure I

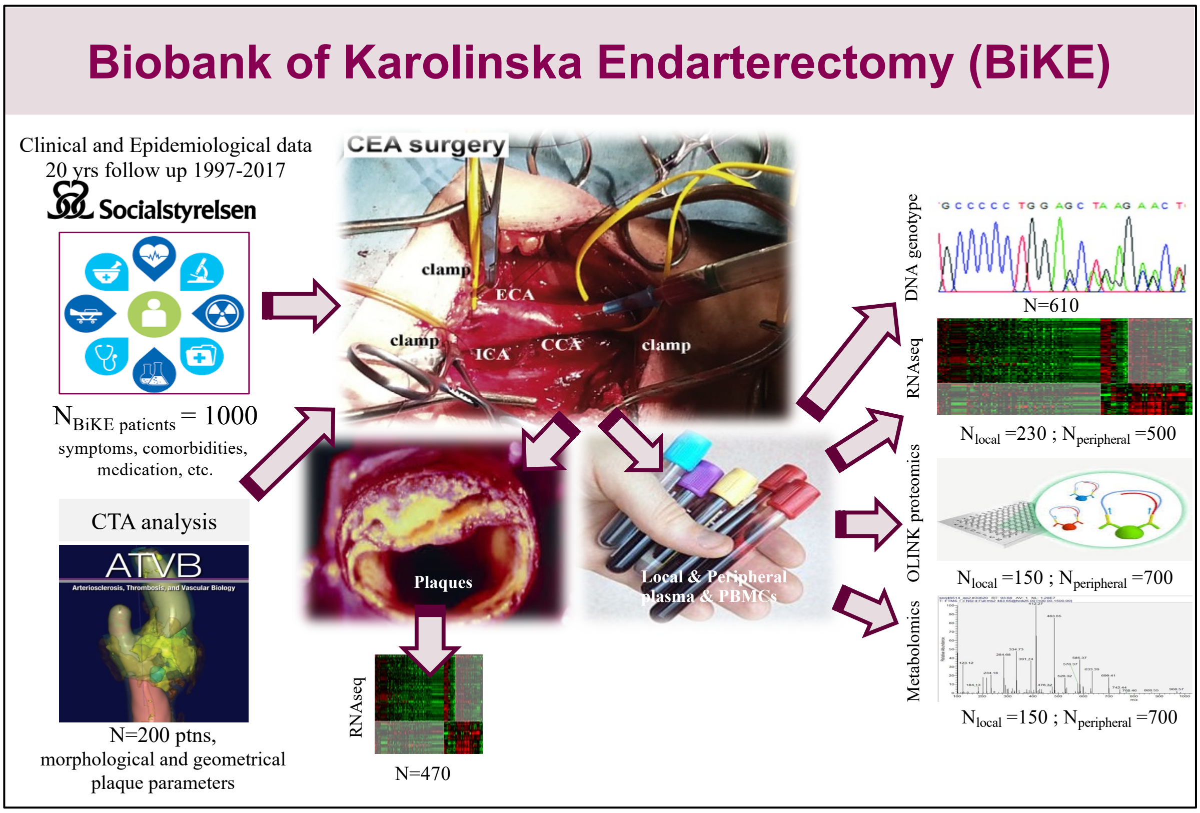


**Supplementary Figure I:** An illustration of all patient samples collected from carotid endarterectomy (CEA) surgery, various omics datasets and clinical/epidemiologic data used in the study. Numbers of individual patient samples profiled with each technology are indicated under the images.

### Supplementary Figure II

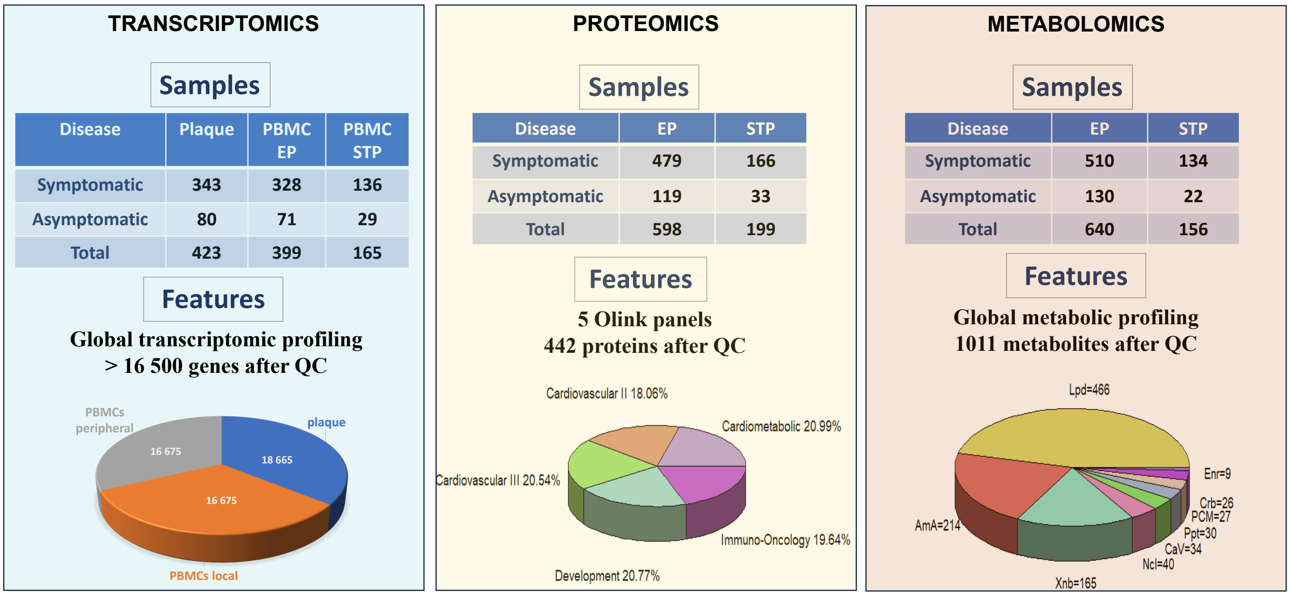


**Supplementary Figure II:** A summary of different samples and analytical features from each layer of single-omics analyses. Abbreviations: PBMC-peripheral blood monocytes, EP-peripheral, STP-local, QC-quality control, Lpd-lipids, AmA-amino acids, Xnb-xenobiotics, Ncl-nucleotides, CaV-cofactors and vitamins, Ppt-peptides, PCM-partially characterized molecules, Crb-carbohydrates, Enr-energy.

### Supplementary Figure III

**TRANSCRIPTOMICS**

**N plaque = 343 S + 80 AS**

**N PBMCs local = 136 S + 29 AS; N PBMCs periph = 328 S + 71 AS**

**A**

**PBMC peripheral**

| Name | P-value | Adjusted p-value | Odds Ratio | Combined score |
|---|---|---|---|---|
| Epithelial Mesenchymal Transition | 1.814e-7 | 0.000009071 | 2.27 | 35.25 |
| TGF-beta Signaling | 0.00003066 | 0.0003833 | 3.32 | 34.55 |
| Mitotic Spindle | 0.000003642 | 0.00007295 | 2.09 | 26.12 |
| G2-M Checkpoint | 0.000004377 | 0.00007295 | 2.07 | 25.54 |
| UV Response Dn | 0.0009642 | 0.009642 | 1.85 | 12.82 |
| Apical Junction | 0.001501 | 0.01251 | 1.66 | 10.79 |
| Wnt-beta Catenin Signaling | 0.01436 | 0.06444 | 2.24 | 9.49 |
| Apoptosis | 0.005269 | 0.03450 | 1.63 | 8.57 |
| Estrogen Response Early | 0.006900 | 0.03450 | 1.53 | 7.64 |
| Myogenesis | 0.006900 | 0.03450 | 1.53 | 7.64 |
| KRAS Signaling Up | 0.006900 | 0.03450 | 1.53 | 7.64 |
| IL-2/STAT5 Signaling | 0.01546 | 0.06444 | 1.46 | 6.10 |
| Androgen Response | 0.03526 | 0.1348 | 1.57 | 5.26 |
| Estrogen Response Late | 0.03774 | 0.1348 | 1.38 | 4.51 |
| Unfolded Protein Response | 0.05224 | 0.1698 | 1.47 | 4.35 |
| Coagulation | 0.05856 | 0.1722 | 1.41 | 3.99 |
| Complement | 0.05435 | 0.1698 | 1.34 | 3.89 |
| PI3K/AKT/mTOR Signaling | 0.09385 | 0.2607 | 1.40 | 3.31 |
| Cholesterol Homeostasis | 0.1190 | 0.3133 | 1.44 | 3.06 |
| Fatty Acid Metabolism | 0.1262 | 0.3154 | 1.27 | 2.63 |
| TNF-alpha Signaling via NF-kB | 0.1405 | 0.3193 | 1.22 | 2.40 |
| mTORC1 Signaling | 0.1405 | 0.3193 | 1.22 | 2.40 |
| Hedgehog Signaling | 0.2008 | 0.3861 | 1.49 | 2.39 |
| Angiogenesis | 0.2008 | 0.3861 | 1.49 | 2.39 |
| p53 Pathway | 0.1839 | 0.3830 | 1.19 | 2.01 |

**PBMC local**

| Name | P-value | Adjusted p-value | Odds Ratio | Combined score |
|---|---|---|---|---|
| Epithelial Mesenchymal Transition | 4.188e-15 | 2.094e-13 | 5.06 | 167.56 |
| Coagulation | 7.298e-11 | 1.825e-9 | 5.01 | 117.01 |
| Complement | 1.345e-7 | 0.000002241 | 3.32 | 52.49 |
| Glycolysis | 0.000001449 | 0.00001811 | 3.05 | 41.06 |
| Xenobiotic Metabolism | 0.000004479 | 0.00004479 | 2.92 | 36.02 |
| Interferon Alpha Response | 0.000008469 | 0.00007058 | 3.96 | 46.28 |
| Interferon Gamma Response | 0.00001328 | 0.00009486 | 2.80 | 31.41 |
| UV Response Dn | 0.00001544 | 0.00009648 | 3.19 | 35.33 |
| TNF-alpha Signaling via NF-kB | 0.00003774 | 0.0001887 | 2.67 | 27.21 |
| Inflammatory Response | 0.00003774 | 0.0001887 | 2.67 | 27.21 |
| Angiogenesis | 0.00006566 | 0.0002984 | 6.18 | 59.56 |
| Cholesterol Homeostasis | 0.00009315 | 0.0003666 | 3.96 | 36.78 |
| Hypoxia | 0.0001026 | 0.0003666 | 2.55 | 23.40 |
| G2-M Checkpoint | 0.0001026 | 0.0003666 | 2.55 | 23.40 |
| IL-2/STAT5 Signaling | 0.0002480 | 0.0007850 | 2.44 | 20.25 |
| Myogenesis | 0.0002669 | 0.0007850 | 2.43 | 19.95 |
| KRAS Signaling Up | 0.0002669 | 0.0007850 | 2.43 | 19.95 |
| heme Metabolism | 0.0006626 | 0.001841 | 2.30 | 16.86 |
| Adipogenesis | 0.001568 | 0.004127 | 2.18 | 14.11 |
| p53 Pathway | 0.003535 | 0.008836 | 2.07 | 11.67 |
| IL-6/JAK/STAT3 Signaling | 0.004982 | 0.01186 | 2.68 | 14.23 |
| E2F Targets | 0.007572 | 0.01721 | 1.95 | 9.53 |
| Apoptosis | 0.009009 | 0.01959 | 2.05 | 9.65 |
| mTORC1 Signaling | 0.02968 | 0.06182 | 1.72 | 6.06 |
| Spermatogenesis | 0.04601 | 0.09202 | 1.81 | 5.56 |

**Plaques**

| Name | P-value | Adjusted p-value | Odds Ratio | Combined score |
|---|---|---|---|---|
| Epithelial Mesenchymal Transition | 6.647e-8 | 0.000003324 | 2.34 | 38.75 |
| TNF-alpha Signaling via NF-kB | 0.000001815 | 0.00004539 | 2.14 | 28.25 |
| Myogenesis | 0.0002477 | 0.003097 | 1.80 | 14.97 |
| Interferon Gamma Response | 0.0002477 | 0.003097 | 1.80 | 14.97 |
| Mitotic Spindle | 0.0003985 | 0.003985 | 1.77 | 13.86 |
| Estrogen Response Early | 0.001436 | 0.01101 | 1.67 | 10.93 |
| Wnt-beta Catenin Signaling | 0.001542 | 0.01101 | 2.84 | 18.40 |
| Hypoxia | 0.002461 | 0.01395 | 1.63 | 9.77 |
| Interferon Alpha Response | 0.002512 | 0.01395 | 1.97 | 11.80 |
| UV Response Dn | 0.003795 | 0.01897 | 1.72 | 9.58 |
| Apical Junction | 0.006717 | 0.02511 | 1.54 | 7.72 |
| mTORC1 Signaling | 0.006717 | 0.02511 | 1.54 | 7.72 |
| Hedgehog Signaling | 0.007031 | 0.02511 | 2.61 | 12.93 |
| Angiogenesis | 0.007031 | 0.02511 | 2.61 | 12.93 |
| G2-M Checkpoint | 0.01070 | 0.03568 | 1.50 | 6.81 |
| IL-6/JAK/STAT3 Signaling | 0.02943 | 0.09198 | 1.66 | 5.85 |
| Androgen Response | 0.04496 | 0.1282 | 1.54 | 4.77 |
| Notch Signaling | 0.04676 | 0.1282 | 2.10 | 6.42 |
| Protein Secretion | 0.04871 | 0.1282 | 1.54 | 4.65 |
| Glycolysis | 0.05406 | 0.1351 | 1.34 | 3.91 |
| Estrogen Response Late | 0.07624 | 0.1815 | 1.30 | 3.35 |
| UV Response Up | 0.09654 | 0.2099 | 1.31 | 3.07 |
| Cholesterol Homeostasis | 0.09883 | 0.2099 | 1.48 | 3.43 |

**B**

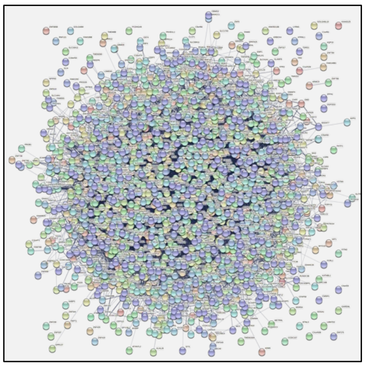

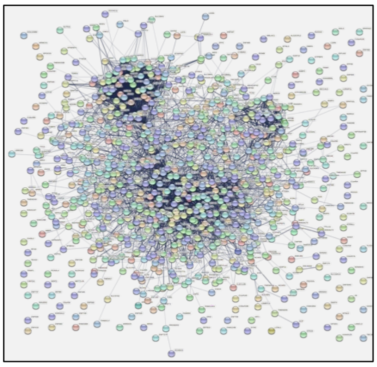

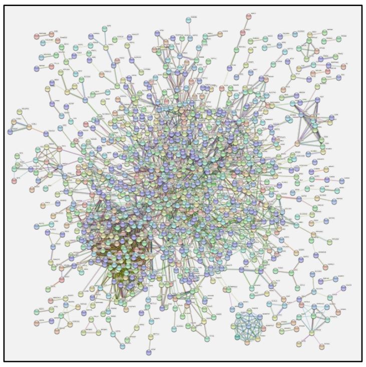

Molecular Function (Gene Ontology) — PBMC peripheral

| GO-term | description | count in network | strength | false discovery rate |
|---|---|---|---|---|
| GO:0005518 | Collagen binding | 22 of 68 | 0.54 | 0.0026 |
| GO:0005201 | Extracellular matrix structural constituent | 29 of 119 | 0.42 | 0.0074 |
| GO:0005178 | Integrin binding | 34 of 147 | 0.4 | 0.0041 |
| GO:0050839 | Cell adhesion molecule binding | 99 of 538 | 0.3 | 4.47e-06 |
| GO:0045296 | Cadherin binding | 58 of 334 | 0.27 | 0.0085 |

Molecular Function (Gene Ontology) — PBMC local

| GO-term | description | count in network | strength | false discovery rate |
|---|---|---|---|---|
| GO:0016638 | Oxidoreductase activity, acting on the ch-n... | 8 of 21 | 0.93 | 0.0142 |
| GO:0043394 | Proteoglycan binding | 9 of 37 | 0.73 | 0.0413 |
| GO:0005518 | Collagen binding | 13 of 68 | 0.63 | 0.0187 |
| GO:0005319 | Lipid transporter activity | 18 of 132 | 0.48 | 0.0326 |
| GO:0051015 | Actin filament binding | 26 of 199 | 0.46 | 0.0082 |

Molecular Function (Gene Ontology) — Plaques

| GO-term | description | count in network | strength | false discovery rate |
|---|---|---|---|---|
| GO:0003779 | Actin binding | 75 of 438 | 0.29 | 0.00047 |
| GO:0008092 | Cytoskeletal protein binding | 136 of 973 | 0.21 | 0.00047 |
| GO:0005515 | Protein binding | 776 of 7026 | 0.1 | 2.05e-11 |
| GO:0043167 | Ion binding | 629 of 6188 | 0.07 | 0.0058 |
| GO:0005488 | Binding | 1211 of 12516 | 0.05 | 4.80e-06 |

**Supplementary Figure III: Transcriptomic data analyses comparing S vs. AS patients. A)** Gene set enrichment analyses using Hallmark database, based on significantly differentially regulated genes from each comparison PBMCs (local and peripheral) or plaques **B)** A mesoscale view of functional associations with enrichment analysis using StringDB, performed on the top 2000 differentially regulated genes (maximum input) from each comparison.

### Supplementary Figure IV

**PROTEOMICS**

**N periph plasma = 479 S + 119 AS**

**N local plasma = 166 S + 33 AS**

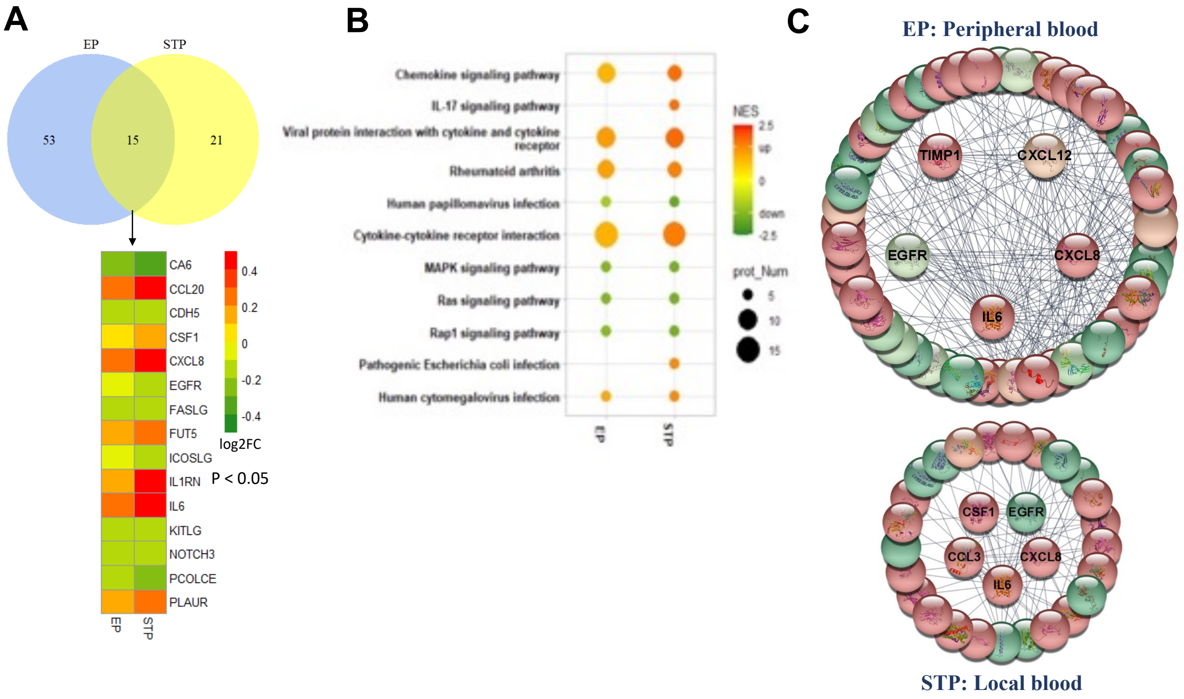


**Supplementary Figure IV: Plasma proteomic data analyses comparing S vs. AS patients. A)** Venn diagram showing overlapping proteins from comparisons of S vs. AS patients' peripheral (EP) or local (STP) plasma. **B)** Functional enrichment analysis of proteins in local and peripheral plasma comparisons. Size of the node illustrates the number of proteins in each pathway. **C)** Evidence-based networks constructed from peripheral and local plasma proteomes comparing S vs. AS patients, with key drivers indicated (red-increased, green-decreased in respective sites).

### Supplementary Figure V

**METABOLOMICS**

**N periph plasma = 510 S + 130 AS**

**N local plasma = 134 S + 22 AS**

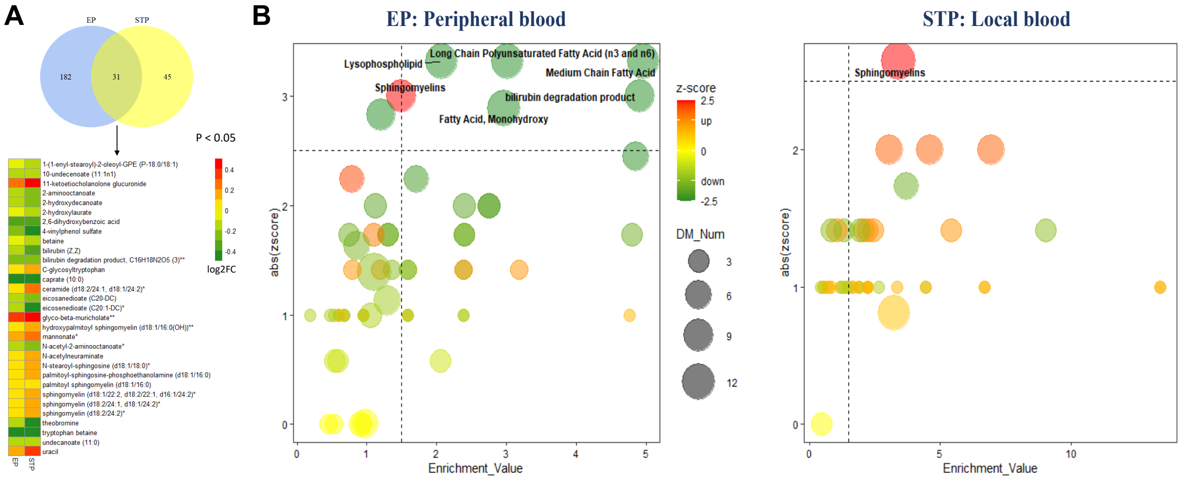


**Supplementary Figure V: Plasma metabolomic data analyses comparing S vs. AS patients. A)** Venn diagram shows overlapping metabolites from plasma analysis comparing S vs. AS patients, both local (STP) and peripheral (EP) blood. **B)** Enrichment analysis of metabolites in local and peripheral blood from S vs AS patients. Size of the node illustrates the number of metabolites in each pathway. Dashed line corresponds to p<0.05.

### Supplementary Figure VI

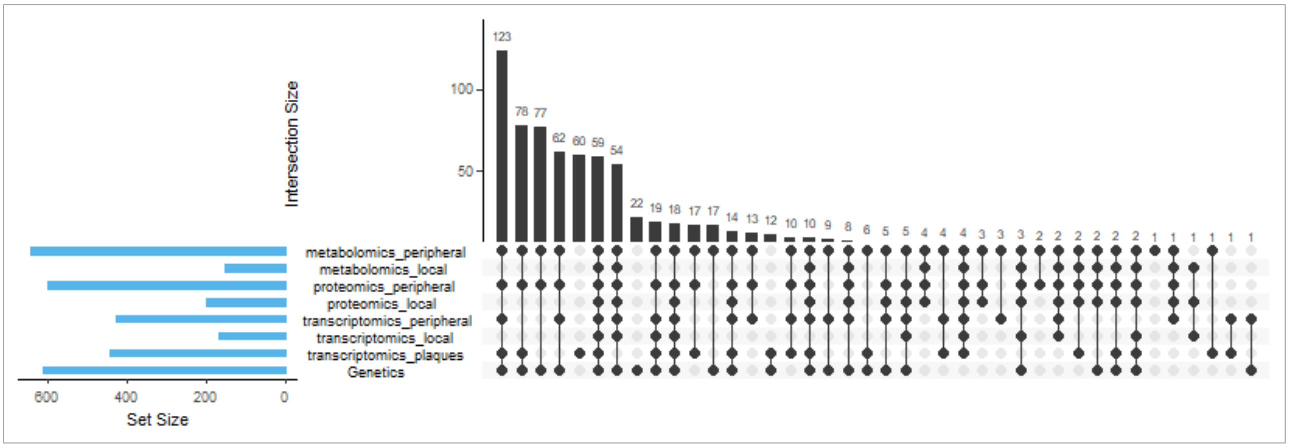


**Supplementary Figure VI:** Overlaps among various omics datasets with numbers of intersection features indicated in blue bar chart to the left, while numbers of patients in intersections are indicated above the black bar chart to the right.

### Supplementary Figure VII

**Combination 1**
**Peripheral site**

| OMICS | Tissue | Feature Numbers | cutoff |
|---|---|---|---|
| Metabolomics | Peripheral plasma | 213 | P < 0.05 |
| Proteomics | Peripheral plasma | 68 | P < 0.05 |
| Transcriptomics | Peripheral PBMCs | 2191 | P < 0.05 |

| Samples | Number |
|---|---|
| Symptomatic | 325 |
| Asymptomatic | 79 |
| Total | 404 |

**Combination 2**
**Local site**

| OMICS | Tissue | Feature Numbers | cutoff |
|---|---|---|---|
| Metabolomics | Local plasma | 1007 | all |
| Proteomics | Local plasma | 442 | all |
| Transcriptomics | Plaque | 3011 | P < 0.05 |

| Samples | Number |
|---|---|
| Symptomatic | 66 |
| Asymptomatic | 13 |
| Total | 79 |

**Combination 3**
**Local + Peripheral**

| OMICS | Tissue | Feature Numbers | cutoff |
|---|---|---|---|
| Metabolomics | Peripheral plasma | 213 | P < 0.05 |
| Proteomics | Peripheral plasma | 68 | P < 0.05 |
| Transcriptomics | Plaque | 3011 | P < 0.05 |

| Samples | Number |
|---|---|
| Symptomatic | 290 |
| Asymptomatic | 68 |
| Total | 358 |

**Supplementary Figure VII:** A summary of different samples and analytical features from each layer of multi-omics analyses.

### Supplementary Figure VIII

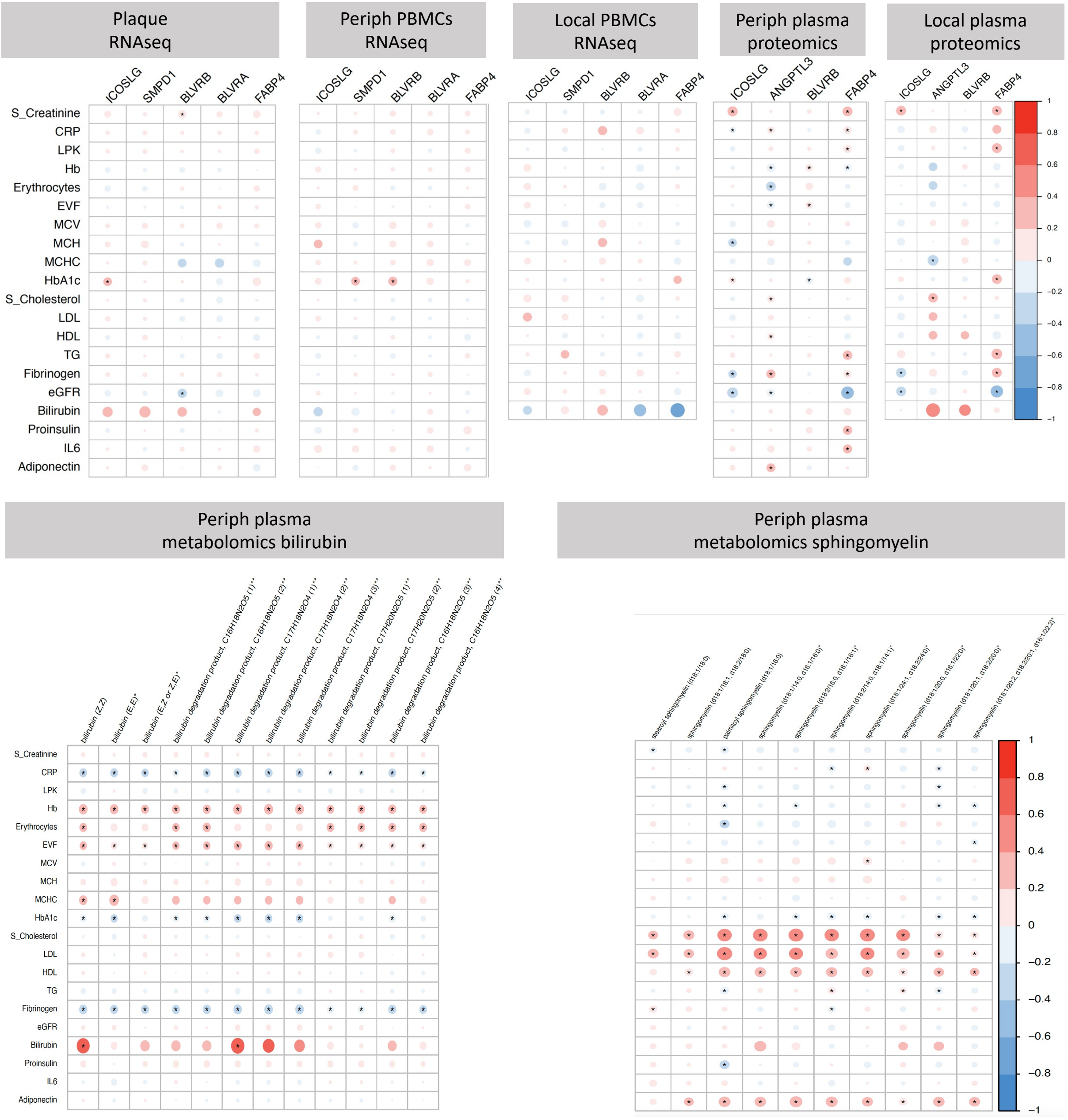


**Supplementary Figure VIII:** Heatmap representation of correlations between transcript, protein or metabolite levels for top targets of interest from omics data and various clinical biochemistry parameters from patients. Correlations were done using Spearman rho method (scale to the right), with correction for multiple comparisons (p-value<0.05 after adjustment indicated with asterisk in the plots). Units for the clinical parameters are as follows: S_Creatinine (serum creatinine, umol/L), CRP (C reactive protein, mg/L), LPK ($10^6$/L), Hb (hemoglobin, %), erythrocytes ($10^{12}$/L), EVF (erythrocyte volume fraction, %), MCV (mean corpuscular volume, fL), MCH (mean corpuscular hemoglobin, pg), MCHC (mean corpuscular hemoglobin concentration, pg), HbA1c (acetylated hemoglobin, mmol/mol), S_Cholesterol (mmol/L), LDL (low density lipoprotein, mmol/L), HDL (high density lipoprotein, mmol/L), TG (triglycerides, mmol/L), Fibrinogen (g/L), eGFR (mL/min/1.73m2), Bilirubin (umol/L), Proinsulin (pmol/L), IL6 (ng/L), Adiponectin (mg/L).

### Supplementary Figure IX

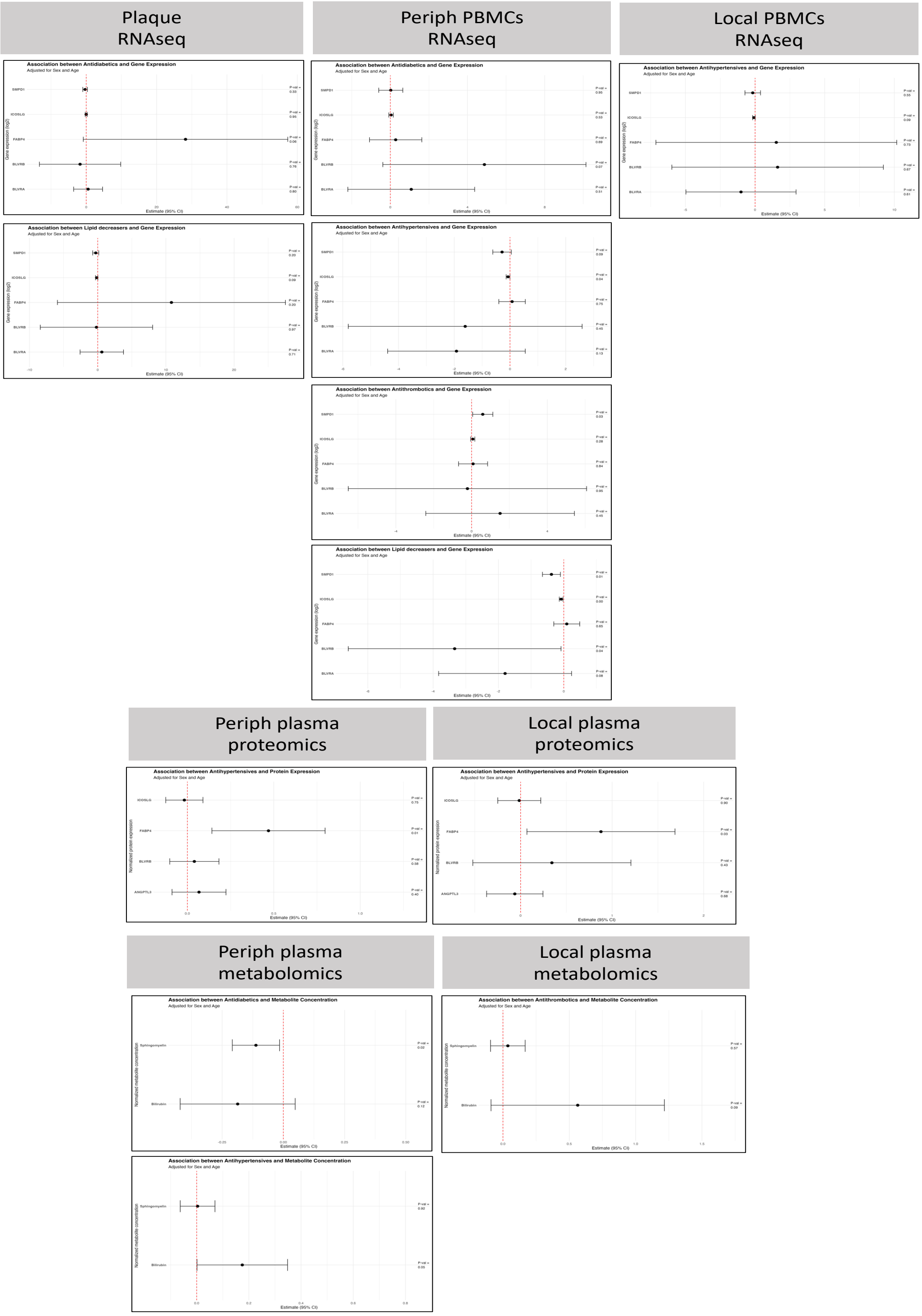


**Supplementary Figure IX:** Forest plot representation of associations between transcript, protein or metabolite levels for top targets of interest from omics data and various medications in the BiKE database. Only selected plots are shown with significant or near-significant results. Confidence intervals (CI) indicated in the x-axis, p-value after adjustment for age and sex indicated in the plots. Medications tested were: Lipid-decreasers (No: 399 patients, Yes: 673), anti-diabetics (No: 363 patients, Yes: 219), anti-hypertensives, and anti-thrombotics (No: 166, Yes: 857).

### Supplementary Figure X

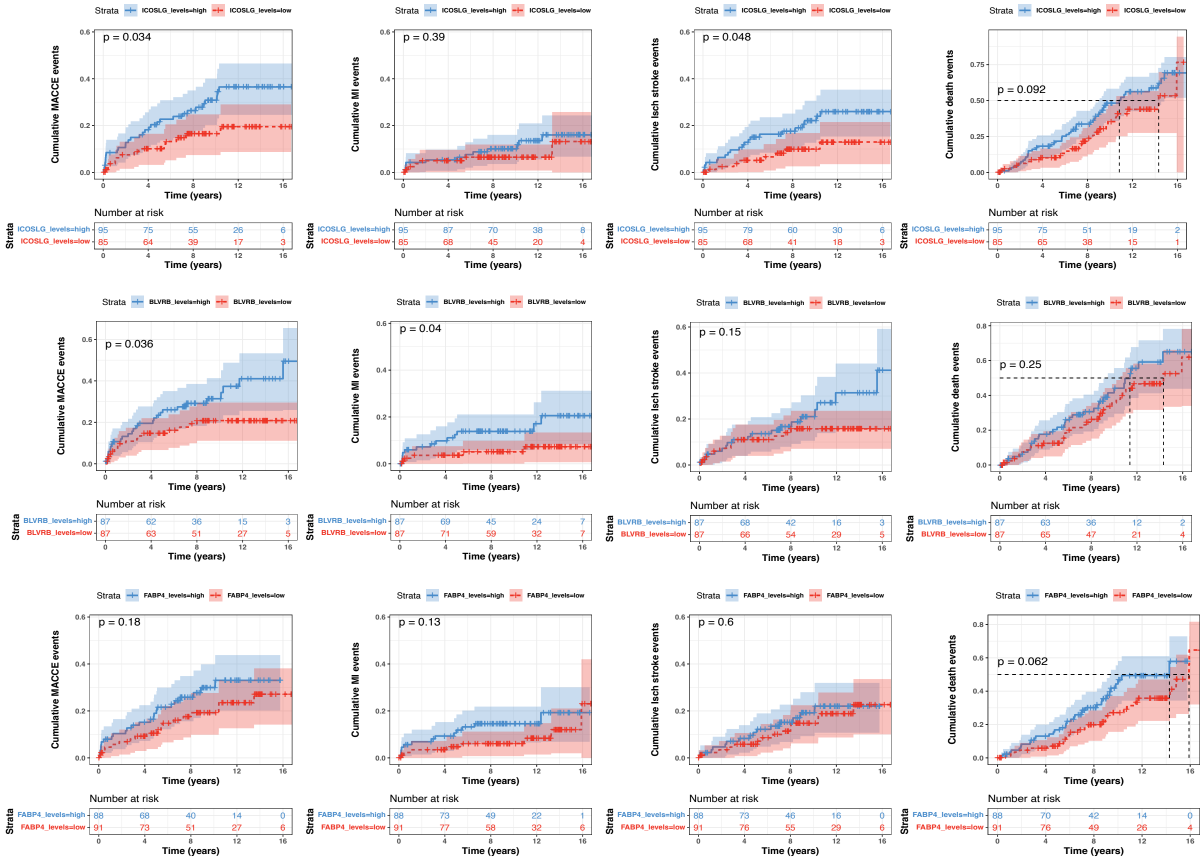


**Supplementary figure X. Cumulative Kaplan-Meier outcome estimates after carotid surgery.** Plots illustrating survival of CEA patients during the 15 years follow-up period after surgery, based on top *vs.* bottom quartile of relevant analyte levels. Both patients that were symptomatic and asymptomatic at surgery were included in the follow-up analysis. Each mark along the lines indicates an event, numbers at risk indicated in tables under the plots. Top raw shows plots for plaque ICOSLG transcript levels. Middle raw shows plots for BLVRB plaque transcript levels, and bottom raw shows FABP4 plaque transcript expression. MACCEs-major adverse cardio- and cerebro-vascular events; MI-myocardial infarction.
